# The death toll of armed conflict and food insecurity in north-east Nigeria, 2016-2019: a statistical study

**DOI:** 10.1101/2022.10.16.22281134

**Authors:** Francesco Checchi, Amy Gimma, Christopher I Jarvis, Kevin van Zandvoort, Abdihamid Warsame

## Abstract

**Background:** Armed conflict, compounded by displacement and food insecurity, has affected Adamawa, Borno and Yobe states of northeast Nigeria (population ≈ 12 million) since 2009. The crisis escalated in 2013-2014 and featured a delayed humanitarian response. We wished to estimate the death rate and toll attributable to the crisis. Data availability constraints restricted estimation to the period 2016 to 2019.

**Methods:** We did a small-area estimation statistical analysis of previously collected data, stratified by local government area and month. We fitted a mixed effects model to household mortality data collected as part of 70 ground surveys implemented by humanitarian actors within the study area and period. Model predictors, selected from a wider candidate list, included livelihood typology, the price of staple cereal, vaccination geo-coverage and the presence of humanitarian actors. To project accurate death tolls, we adjusted population denominators based on internal and refugee displacements. We used the model and population estimates to project mortality under observed conditions and under different assumptions of counterfactual conditions, had there been no crisis, with the difference between observed and counterfactual providing excess mortality.

**Results:** Crude and under 5 years death rates were highly elevated across most ground surveys, with net negative household migration. Between April 2016 and December 2019, we projected that 490,000 deaths (230,000 children under 5y) died in excess of the counterfactual non-crisis level; this death toll ranged from medians of 90,000 to 550,000 depending on the counterfactual assumptions chosen, specifically whether to consider a Nigeria-wide price inflation phase as inherently part of the crisis in the north-east. Crude and under 5 years death rates were two to three times higher than counterfactual levels, and highest in 2016-2017.

**Discussion:** This may be the first crisis-wide estimate of mortality attributable to the crisis in north-east Nigeria. Our findings do not reflect acute emergency periods before 2016 and the situation in neighbouring countries affected by the conflict. Sensitivity analysis suggests that results for Borno state, where conflict has been most intense, are subject to under-estimation due to data not representing hard-to-access areas. Further studies to document mortality in this and other crises are needed to guide decision-making and memorialise their human toll.

## Introduction

### The crisis in northeast Nigeria

A large region of north-east Nigeria and, increasingly, neighbouring countries including Nigeria, Chad and Cameroon, have been affected by protracted crisis conditions linked to an armed conflict between authorities, local defence units and the Boko Haram group. Poverty, inequality and environmental shocks (including the loss of livelihoods around Lake Chad) are considered root causes of the conflict [1–3]. Within Nigeria, Adamawa, Borno and Yobe states have been disproportionately affected. Insecurity escalated in 2013 and 2014, leaving 1.7M people internally displaced by mid-2015, a ten-fold increase over two years [4].

The armed conflict has been marked by widely reported attacks against civilians, including systematic sexual violence, arbitrary detention and damage to some two thirds of health facilities in the region [5]. International humanitarian assistance remained minimal until 2016 [6]. The response has been heavily securitised, with aid actors largely prevented from operating outside of military-controlled enclaves hosting large concentrations of displaced or forcibly relocated people [7]. The situation has been chronically alarming in inaccessible areas, where some 1.2M civilians were estimated to remain as of end 2019, out of some 7.9M in need across Adamawa, Borno and Yobe [4].

During 2016-2018, loss of livelihoods and access to functional markets due to the crisis combined with countrywide inflation-related food price increases to greatly exacerbate pre-existing food insecurity [8]. As of mid-2016, about 34% of the three states’ population was projected to be in phases 3 to 5 of the Integrated Phase Classification of food and nutrition insecurity, with suspected pockets of famine [9, 10]. Measles and cholera epidemics, common manifestations of forced displacement and acute malnutrition, also occurred [11].

### Scope of this study

Armed conflict and food insecurity, especially when protracted, are characterised by increased population mortality, both directly (violence) and indirectly (increased risk of disease, reduced access to healthcare) attributable to the crisis [12]. Information on mortality can inform the ongoing humanitarian response by providing a measure of the gap in avertable deaths remaining to be filled, provide evidence for resource mobilisation, and support conflict resolution [13]. We sought to quantify mortality attributable to crisis conditions in Borno, Adamawa and Yobe states from 2016 to 2019. The analysis was finalised with a year’s delay due to the COVID-19 pandemic.

## Methods

### Overview of the approach

We used a small-area estimation [14] statistical modelling approach relying on datasets collected by various organisations as part of the humanitarian response in Northeast Nigeria, and not requiring primary data collection. The method consists of six steps, extensively described in a separate paper [15] and enables estimation by Local Government Area or LGA (equivalent to a district or county) and month. Statistical code and datasets to implement the analysis using R software [16] are available on https://github.com/francescochecchi/mortality_small_area_estimation/tree/nga: these allow users to vary various data management and modelling parameters by modifying their values on Microsoft Excel input files.

Briefly, the sequential steps comprise of (1) identifying, managing and grading the quality of existing population mortality surveys done in the region and time period of interest (these surveys provide the data on which the model is trained); (2) reconstructing population denominators for each LGA-month by applying assumed growth rates to existing alternative point-in-time population source, while also adjusting for internal and refugee displacement in and out of each LGA and grading the quality of each alternative source to compute a weighted mean (population denominators are used to convert between death rates and tolls and to calculate per capita quantities of model predictors); (3) identifying datasets that offer proxy or direct measures of various potential factors along a causal pathway leading to mortality, and preparing these potential “predictor” datasets for analysis by imputing missing values, smoothing or interpolation, creating lags, etc.; (4) combining ground mortality, population and predictor data into a statistical model, selected on the basis of predictive accuracy on cross-validation (i.e. on a simulated external dataset), the plausibility of observed associations and the sensitivity of predictor variables to crisis conditions; (5) using the model to predict mortality (both death toll and rate) for all LGA-months, (i) as per the observed data values and (ii) per counterfactual scenarios defined by assumptions about which values the model predictors and population denominators might have taken in the absence of a crisis – here, the difference between observed-conditions and counterfactual predictions provides the estimated excess mortality; and (6) performing specific sensitivity analyses, e.g. to explore the effect of potential bias in input data. We implement steps (4) to (6) for both the crude death rate (CDR, deaths due to all causes among all ages per person-time) and the under 5 years death rate (U5DR, deaths due to all causes among children under 5y per child-time). Step (2) does not depend on the others steps and its methods are specific to the Nigeria context: for brevity’s sake this step is summarised in the Supplementary Material.

### Study population and timeframe

We collected data up to December 2019; we restricted the analysis period to 2016-2019 due to data sparsity pre-2016 and the unclear effect of the COVID-19 pandemic in 2020. However, pre-2016 values for predictor variables, where available, were used to inform counterfactual scenarios. Adamawa, Borno and Yobe states are divided into 21, 27 and 17 LGAs respectively, and, as of July 2017 (the mid-point of our analysis period), had populations of 3.9 (8% internally displaced), 5.0 (29%) and 3.4 (11%) million, namely 12.3 million overall (18%), according to our reconstruction (see Supplementary Material).

### Data sources

#### Ground mortality data

We accessed reports and raw datasets of 73 multi-stage cluster sampling household surveys conducted in the region during 2016-2018 by either the National Bureau of Statistics (NBS, supported by Unicef and the United States Centers for Disease Control and Prevention; N = 57), Unicef (N = 7) or Action Against Hunger (N = 9), all using the standardised methods, survey instruments and analytic tools of the Standardised Monitoring and Assessment of Relief and Transition (SMART) project [17]. SMART surveys are routinely conducted across crisis settings; while primarily focussing on nutritional status, they usually feature a mortality questionnaire module that elicits information from respondents on the size and evolution (births, deaths, arrivals and departures) of their nuclear (i.e. regularly sharing meals) household members over a retrospective ‘recall’ period, enabling estimation of various demographic quantities including the CDR and U5DR [18]. The sampling universe of NBS surveys was a ‘domain’ (grouping of LGAs: Supplementary Material), but raw datasets contained information on which LGA each survey cluster fell within; remaining surveys were conducted with single LGAs as their sampling universe. Surveys were implemented at regular intervals with a median recall period of 135 days (range 90 to 309): the combination of frequency and geographic breadth of data collection yielded high person-time survey coverage (Supplementary Material). We excluded two surveys due to incomplete datasets and one due to highly unusual estimates. For each of the remaining 70, we re-analysed raw datasets, extracted quality scores (Supplementary Material) and reviewed the report to quantify possible selection bias (59/70 surveys reportedly sampled only a fraction from 8% to 97% of the intended sampling universe).

#### Predictor data

Guided by an *a priori* causal framework [12], we identified candidate datasets from which to attain information on predictor variables (Table 1) through internet searches and meetings with governmental agencies and humanitarian coordination mechanisms in Maiduguri and Abuja. Details on data management are in the Supplementary Material. Additional predictors of interest had incomplete geographic (e.g. only Borno state) or time period coverage, or were too sparse to support imputation: these included tracking of attacks against aid workers; settlement intactness and habitation data collected by the polio eradication programme; Emergency Food Security Assessments done at LGA, domain or state level once or twice a year; health facility functionality data collected through the Health Resources Availability Monitoring System (HeRAMS) by the World Health Organisation; epidemic surveillance data from WHO’s Early Warning, Alert and Response System (EWARS) in Borno; and measles incidence data from Yobe’s State Ministry of Health.

**Table 1.** Candidate predictor datasets used to build the model. With the exception of market prices (see Supplementary Material), all variables were available at the LGA-month level of stratification.

| Variable | Value(s) | Source and URL if public | Notes and assumptions |
| --- | --- | --- | --- |
| **Climate** |  |  |  |
| Rainfall | Mean of Standard Precipitation Index (number of standard deviations away from the historical average rainfall during the same period of the year) | Climate Engine https://clim-engine-development.appspot.com/fewsNet | Used 5-day Climate Hazards Group InfraRed Precipitation with Station data (CHIRPS) dataset [19], with historical data starting in 1981. Explored lags of 0-6mths and 2-3mth running means. |
| Season | Whether a given month is in the lean season or harvest season | Famine Early Warning Systems Network http://fews.net/file/87830 | As per expectations of a typical year. |
| **Insecurity** |  |  |  |
| Incidence of insecurity events† | events per 100,000 population | Armed Conflict Location & Event Data Project (ACLED) [20] https://acleddata.com | Retained all ACLED event types. Explored lags of 0-4mths. |
| Rate of people killed† | deaths per 100,000 population |  |  |
| Rate of people killed† | deaths per 100,000 population | Nigeria Security Tracker, Council on Foreign Relations https://www.cfr.org/nigeria/nigeria-security-tracker/p29483 | Explored lags of 0-4mths. |
| Incidence of insecurity events† | events per 100,000 population | Global Terrorism Database, University of Maryland https://www.start.umd.edu/gtd/ | The database is restricted to violence by non-state actors. We used reported GPS coordinates to identify the LGA. Explored lags of 0-4mths. |
| Rate of people killed† | deaths per 100,000 population |  |  |
| Incidence of insecurity events† | events per 100,000 population | Nigeria Watch, Institut de Recherche pour le Développement http://www.nigeriawatch.org/ (available at a fee) | Included all events tagged as 'political issue', 'religious issue', 'land issue', 'market issue' and 'cattle grazing'. Explored lags of 0-4mths. |
| Rate of people killed† | deaths per 100,000 population |  |  |
| **Displacement** |  |  |  |
| Proportion of IDPs† | IDPs living within the LGA, out of LGA population | As per our population estimation. |  |
| IDP departure rate† | IDPs leaving the LGA per 100,000 | As per our population estimation. |  |
| IDP arrival rate† | IDPs arriving into the LGA per 100,000 | As per our population estimation. |  |
| **Food security and livelihoods** |  |  |  |
| Main local livelihood type | Pastoralists, agro-pastoralists | Famine Early Warning Systems Network http://fews.net/west-africa/nigeria/livelihood-zone-map/february-2019 | Grouped 11 livelihood zones into two categories. Assumed to be static. |
| Market prices | Price of 1 Kg of staple cereals (wholesale or retail)<br>Price of 1 L of gasoline fuel<br>Price of 1 loaf of bread | World Food Programme https://data.humdata.org/dataset/wfp-food-prices-for-nigeria | Explored lags of 0-3mths. See Supplementary Material for further notes on data management. |
| **Humanitarian (public health) services** |  |  |  |
| Humanitarian actor presence† | Number of humanitarian actors present per 100,000 population (all sectors; public health only = health, nutrition and water, hygiene & sanitation) | United Nations Office for Coordination of Humanitarian Affairs https://reliefweb.int/updates?advanced-search=%28C175%29_%28T4590%29_%28F12570%29 | Explored lags of 0-3mths. |
| Accessibility by humanitarian actors | Partial / none, Full | Reconstructed by the authors based on systematic review of UN infographics and maps https://reliefweb.int/updates?view=maps&advanced-search=%28C175%29_%28F12570%29 |  |
| Incidence of admissions for severe acute malnutrition† | Number of children admitted to therapeutic feeding services per 100,000 population | Nutrition Cluster Information Working Group, Abuja / Maiduguri (not public) | Proxy for nutritional service coverage. Also a proxy for nutritional status. |
| Quality of case management of severe acute malnutrition | Proportion of children admitted who are discharged successfully (survive and are not lost to follow-up or transferred) | Nutrition Cluster Information Working Group, Abuja / Maiduguri (not public) | Proxy for service quality. |
| Vaccination geo-coverage |  | Nigeria Vaccination Tracking System http://vts.eocng.org/ChronicalCoverage/Index (website no longer accessible) [21–23] | Proxy for overall health service availability and coverage. See Supplementary Material for further notes on data management. |

† Divided by district population estimates to obtain a population rate.

### Analysis

#### Predictive model

Given the data structure (household-level mortality observations; LGA-level predictor data; multiple rounds of mortality surveys for each LGA, with *de novo* samples each time), we fitted a mixed quasi-Poisson model to the count of deaths within each household, offset by the natural log of household person-time at risk during the survey recall period, with a random effect for LGA and weights for survey robustness scores (range 0 to 1; see Supplementary Material). We calculated mean monthly values of analysis-eligible predictors over the recall period of each mortality survey. For both CDR and U5DR, we selected predictors by (i) searching across all plausible lags and continuous versus categorical versions of each variable; (ii) combining the most data-consistent (lowest F-test p-value) among these variants into all possible multivariable models, and shortlisting models based on predictive accuracy (low Dawid-Sebastiani scoring rule [24]); (iii) selecting a single fixed-effects model from the shortlist based on its performance on 10-fold LGA-level cross-validation and the plausibility of observed associations; (iv) testing for plausible interactions; and (v) adding the random effect, retaining the mixed model if it offered superior predictive accuracy on cross-validation. We used the g 1mmTMB package for mixed modelling [25].

#### Mortality estimation

Excess mortality may be defined as the difference between deaths under observed and counterfactual conditions, the latter constituting expected levels in the absence of a crisis. We thus defined central (i.e. average), reasonable best- and reasonable worst-case counterfactual scenarios for predictors and population denominators (Table 2). We used the model to predict mortality given the observed and counterfactual data, and the resulting excess. We computed bootstrapping confidence intervals by resampling from the predictions’ standard errors.

**Table 2.** Counterfactual value assumptions.

| Variable | Scenario | Value | Justification |
| --- | --- | --- | --- |
| Predictors |  |  |  |
| Price of white maize | central | Mean of values in Jan-Jun 2015 and 2018-2020, for each LGA | Available periods before and after the price inflation period, as determined through visual inspection (see Supplementary Material). |
|  | worst | Mean of entire 2015-2020 series for each LGA | Assumes that periods of price inflation do not constitute an unusual crisis, but rather are part of normal fluctuation in prices. |
|  | best | Minimum of entire 2015-2020 series for each LGA | Most optimistic assumption given available data. |
| Presence of humanitarian actors | central | Baseline at the start (generally very limited humanitarian presence) | It is unlikely that humanitarian actors would have expanded their presence in the absence of armed conflict. |
|  | worst | No humanitarian assistance whatsoever |  |
|  | best | Baseline at the start (generally very limited humanitarian presence) | As above. |
| Vaccination geo-coverage | central | Point estimate of coverage in non-BAY† states, by month in 2016-2019 | In the absence of armed conflict, the affected region would have achieved a coverage no worse than elsewhere in Nigeria. See Supplementary Material for data sources. |
|  | worst | lower 95%CI of coverage in non-BAY states, by month in 2016-2019 |  |
|  | best | upper 95%CI of coverage in non-BAY states, by month in 2016-2019 |  |
| Population denominators |  |  |  |
| Number of IDPs | central | average proportion of displacement not due to the insurgency (= 5%, from IOM dataset) | Some displacement did occur specifically due to natural disasters. |
|  | worst | 20% of the actual level |  |
|  | best | no displacement |  |
| Number of refugees | central | average proportion of displacement not due to the insurgency (= 5%, from IOM dataset) | As above. |
|  | worst | 20% of the actual level |  |
|  | best | no displacement |  |

† BAY = Borno, Adamawa, Yobe.

### Ethics

All data were previously collected for routine humanitarian response and/or health service provision purposes, and were either in the public domain or shared in fully anonymised format. The study was approved by the Ethics Committee of the London School of Hygiene & Tropical Medicine (ref. 15334) and the Nigerian Institute of Medical Research Institutional Review Board (ref. IRB/18/065).

## Results

### Crude mortality patterns

Re-analysis of available SMART surveys (Table 3) showed that CDR (median 0.55 per 10,000 person-days or 20 per 1000 person-years) was generally elevated, particularly in 2017, compared to the projected median for Nigeria during 2015-2020 (0.34 per 10,000 person-days or 12 per 1000 person-years) [26]. Both CDR and U5DR reached peaks consistent with catastrophic emergencies [27]. Injury appeared to be a leading cause of death. While in Nigeria as a whole the ratio of infant to under 5y mortality was 62/102 per 1000 live births [26], in Adamawa, Borno and Yobe surveys suggested a lower proportion of infants among under 5y deaths. Crude birth rate was slightly lower than nationally (38 per 1000 person-years), and surveys mostly reported negative net household migration. Generally, the most extreme estimates were recorded in late 2016 to middle 2017 (Figure 1).

**Table 3.** Characteristics of analysis-eligible ground mortality surveys. The median (inter-quartile range) of survey point estimates are reported unless noted.

|  | Overall | 2016 | 2017 | 2018 |
| --- | --- | --- | --- | --- |
| Eligible surveys (N) | 70 | 13 | 31 | 26 |
| Crude death rate<br>(per 10,000 person-days) | 0.55<br>(0.17 to 1.58) | 0.48<br>(0.25 to 1.58) | 0.83<br>(0.29 to 1.57) | 0.46<br>(0.17 to 1.06) |
| Under 5 years death rate<br>(per 10,000 child-days) | 1.14<br>(0.23 to 4.46) | 0.94<br>(0.53 to 1.90) | 1.75<br>(0.56 to 4.46) | 0.79<br>(0.23 to 2.25) |
| Proportion of <5yo deaths that<br>were among infants <1yo | 0.35<br>(0.00 to 1.00) | 0.29<br>(0.00 to 0.60) | 0.35<br>(0.00 to 0.80) | 0.38<br>(0.00 to 1.00) |
| Household size (n) | 5.3<br>(3.8 to 7.3) | 5.3<br>(3.8 to 6.0) | 5.3<br>(4.3 to 6.3) | 5.3<br>(4.4 to 7.3) |
| Proportion of children aged<br>under 5y | 0.19<br>(0.14 to 0.29) | 0.18<br>(0.14 to 0.29) | 0.19<br>(0.16 to 0.26) | 0.18<br>(0.16 to 0.22) |
| Proportion of females in<br>household | 0.50<br>(0.47 to 0.60) | 0.51<br>(0.50 to 0.60) | 0.50<br>(0.47 to 0.53) | 0.50<br>(0.48 to 0.52) |
| Crude birth rate<br>(per 1000 person-years) | 33.7<br>(7.3 to 90.3) | 37.4<br>(17.9 to 44.6) | 41.7<br>(14.2 to 82.7) | 23.7<br>(7.3 to 90.3) |
| Net migration rate<br>(per 1000 person-years) | -42.0<br>(-119.8 to 133.3) | -8.1<br>(-52.3 to 133.3) | -63.2<br>(-119.8 to 0.0) | -48.7<br>(-89.9 to -5.2) |
| Injury-specific death rate†<br>(per 10,000 person-days) | 0.16<br>(0.00 to 1.44) | 1.07<br>(0.28 to 1.44) | 0.02<br>(0.00 to 0.78) | no data |

† Based on 25 surveys that collected this information (5 in 2016, 20 in 2017 and none in 2018).

**Figure 1.**
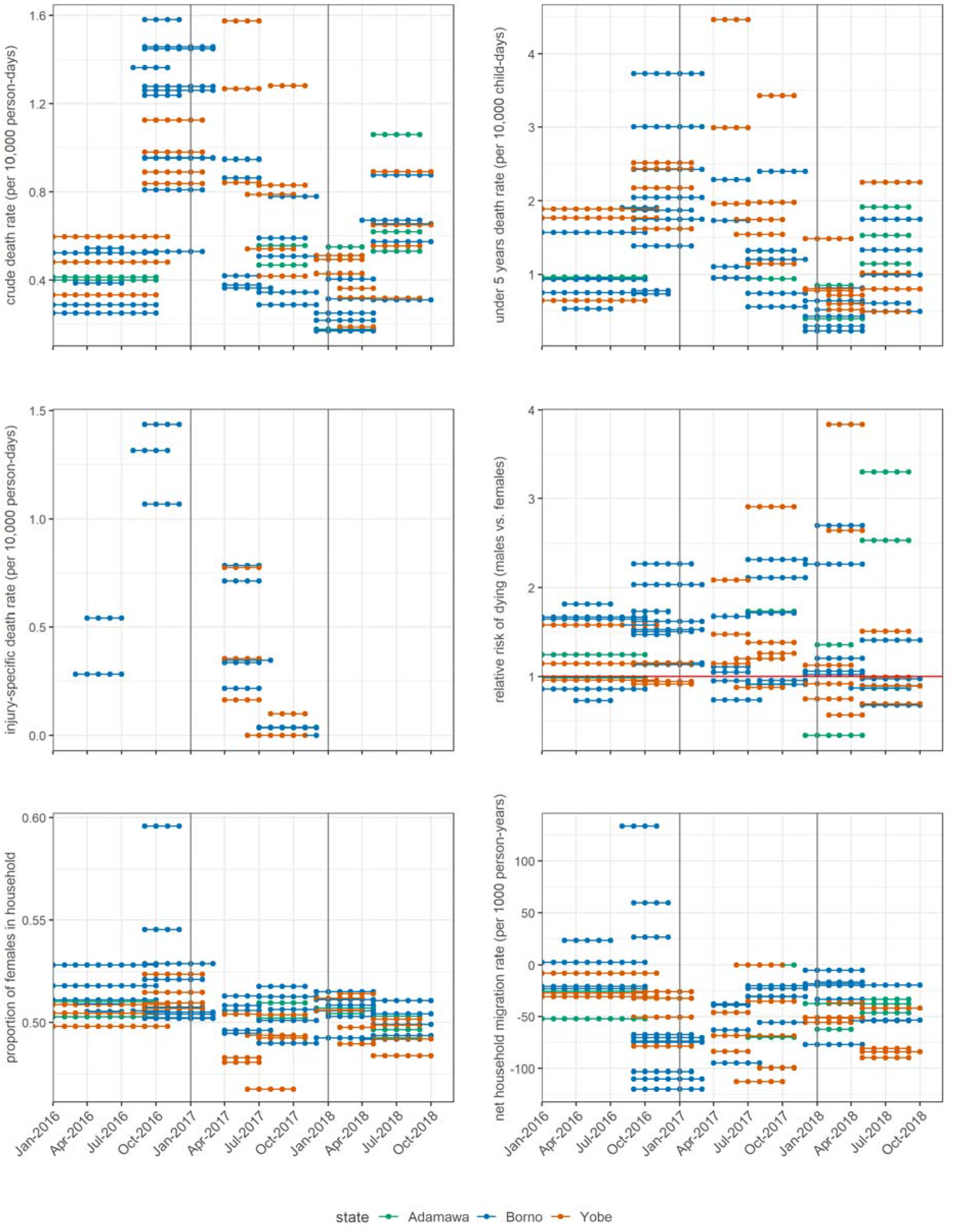
Trends in key survey-estimated demographic indicators. Each dotted segment represents the recall period of a single survey.

### Predictive model

The final models for CDR and U5DR consisted of the same four predictor variables (Table 4). CDR was higher among agro-pastoralists and with increasing price of maize, one of the main cereal staples in northeast Nigeria; it decreased as humanitarian actor presence intensified, and with increasing vaccination geo-coverage. For U5DR, the association with geo-coverage was not evident. Model predictions at the LGA level were mostly well aligned with observations (Supplementary Material), with minor systematic underprediction (Table 4).

**Table 4.** Fixed-effect model coefficients and measures of predictive performance.

| Fixed effect | Crude death rate |  | Under 5y death rate |  |
| --- | --- | --- | --- | --- |
|  | Rate ratio (95%CI)<sup>†</sup> | p-value | Rate ratio (95%CI)<sup>†</sup> | p-value |
| Main livelihood type |  |  |  |  |
| Pastoralists [ref.] | - |  | - |  |
| Agro-pastoralists | 1.20 (0.97 to 1.49) | 0.098 | 1.29 (1.01 to 1.66) | 0.043 |
| Number of humanitarian actors per 100,000 population (lag = 3mths) |  |  |  |  |
| ≤ 2.5 [ref.] | - |  | - |  |
| 2.5 to 4.9 | 0.99 (0.82 to 1.19) | 0.905 | 0.95 (0.74 to 1.23) | 0.702 |
| 5.0 to 7.4 | 0.87 (0.68 to 1.11) | 0.257 | 0.99 (0.71 to 1.37) | 0.954 |
| ≥ 7.5 | 0.61 (0.46 to 0.81) | < 0.001 | 0.67 (0.48 to 0.94) | 0.022 |
| Wholesale price of 1Kg of white maize (USD, 2010) (lag = 3mths) |  |  |  |  |
| ≤ 0.25 [ref.] | - |  | - |  |
| 0.25-0.34 | 1.50 (1.12 to 2.02) | 0.007 | 1.65 (1.04 to 2.62) | 0.032 |
| ≥ 0.35 | 2.29 (1.73 to 3.03) | < 0.001 | 2.93 (1.91 to 4.50) | < 0.001 |
| Vaccination geo-coverage |  |  |  |  |
| ≤ 25% [ref.] | - |  | - |  |
| 25% to 49% | 1.03 (0.62 to 1.70) | 0.917 | 1.35 (0.56 to 3.22) | 0.500 |
| 50% to 74% | 0.82 (0.49 to 1.36) | 0.444 | 1.26 (0.53 to 3.00) | 0.597 |
| ≥ 75% | 0.50 (0.24 to 1.01) | 0.052 | 0.68 (0.23 to 2.03) | 0.486 |
| Predictive performance (at the LGA level) |  |  |  |  |
| Dawid-Sebastiani score (on cross-validation) | 32.6 (214.9) |  | 74.3 (134.4) |  |
| Mean square error (on cross-validation) | 51.3 (252.1) |  | 15.8 (41.6) |  |
| Relative bias | -8.7% |  | -7.1% |  |
| Relative precision of 95%CI<sup>†</sup> | ±59.7% |  | ±60.1% |  |

<sup>†</sup>CI = Confidence Interval.

### Estimated mortality

Over a 45-month period from April 2016 to December 2019, we estimate that about 910,000 deaths occurred in the three states combined (41% among children under 5y; Table 5). The central counterfactual scenario yielded an excess death toll of 490,000 (47% children under 5y); however, in the reasonable worst-case counterfactual scenario (i.e. highest non-crisis death rate leading to the best-case excess scenario), a far smaller crisis-attributable death toll (90,000) was projected.

**Table 5.**
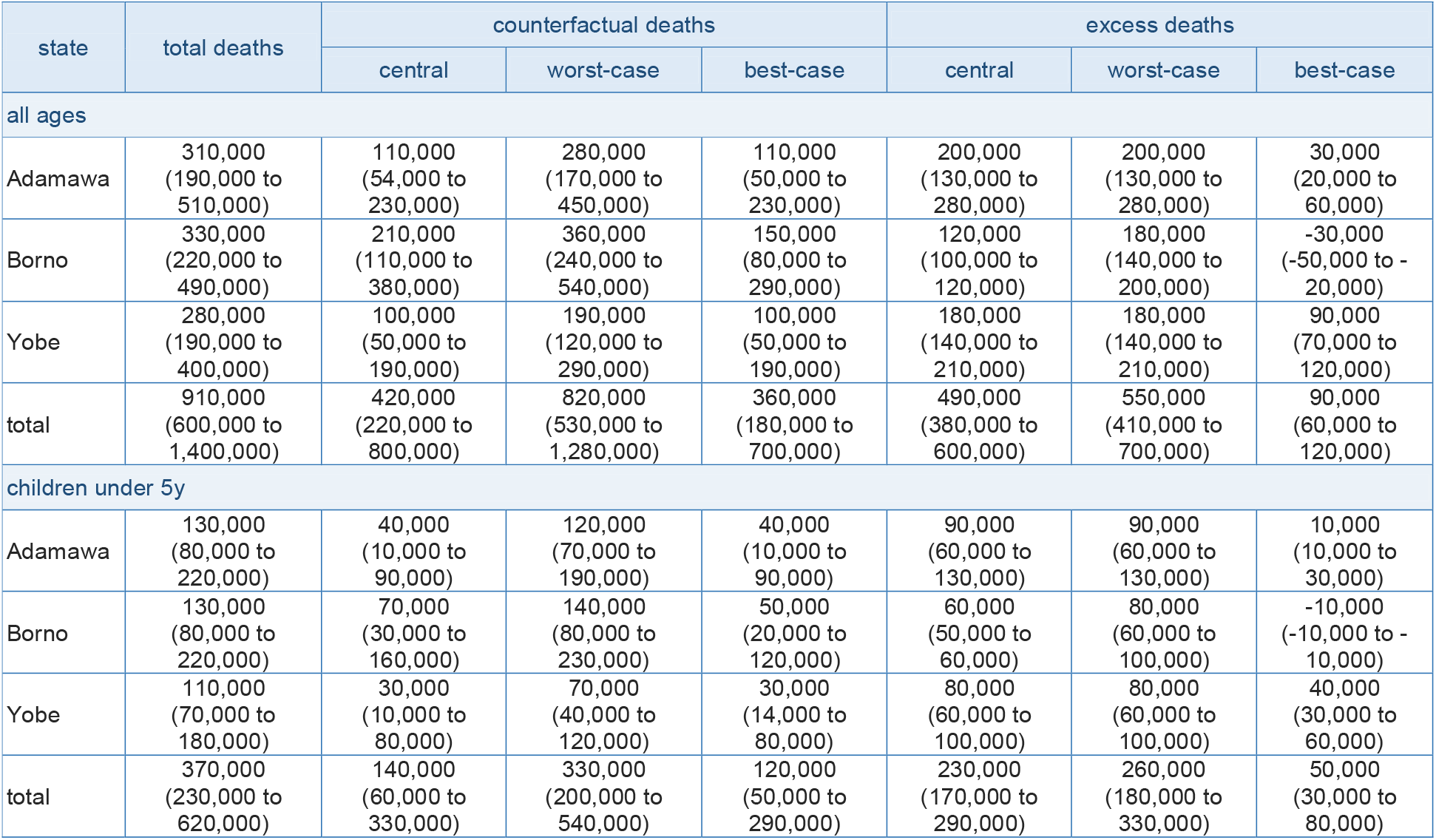
Estimates (95% confidence intervals) of actual, counterfactual and excess mortality, for all ages and children under 5y old, by state and counterfactual scenario.

The highest death tolls occurred in 2016 and 2017, with more moderate levels in 2018 and 2019 (Table 6). CDR and U5DR estimates across the region displayed a similar trend (Figure 2). On average, CDR and U5DR were between two and three times higher than the likely counterfactual levels. Note that the differences between actual and counterfactual death rates are not linearly related to the differences in death tolls, as counterfactual scenarios also vary the extent of displacement and thus the denominators at risk across different LGAs.

**Table 6.** Estimates (95% confidence intervals) of total and excess mortality, for all ages and children under 5y old, by year. Only results for the most likely counterfactual scenario are shown.

| year | all ages |  | children under 5y |  |
| --- | --- | --- | --- | --- |
|  | total deaths | excess deaths | total deaths | excess deaths |
| 2016† | 240,000<br>(160,000 to 370,000) | 160,000<br>(120,000 to 210,000) | 100,000<br>(60,000 to 160,000) | 70,000<br>(50,000 to 100,000) |
| 2017 | 320,000<br>(210,000 to 480,000) | 210,000<br>(160,000 to 270,000) | 140,000<br>(90,000 to 230,000) | 110,000<br>(70,000 to 140,000) |
| 2018 | 190,000<br>(130,000 to 300,000) | 80,000<br>(70,000 to 90,000) | 70,000<br>(40,000 to 130,000) | 40,000<br>(30,000 to 40,000) |
| 2019 | 160,000<br>(100,000 to 250,000) | 40,000<br>(30,000 to 40,000) | 60,000<br>(30,000 to 100,000) | 20,000<br>(1,000 to 20,000) |

† Starting from April 2016, i.e. only 9 months.

**Figure 2.**
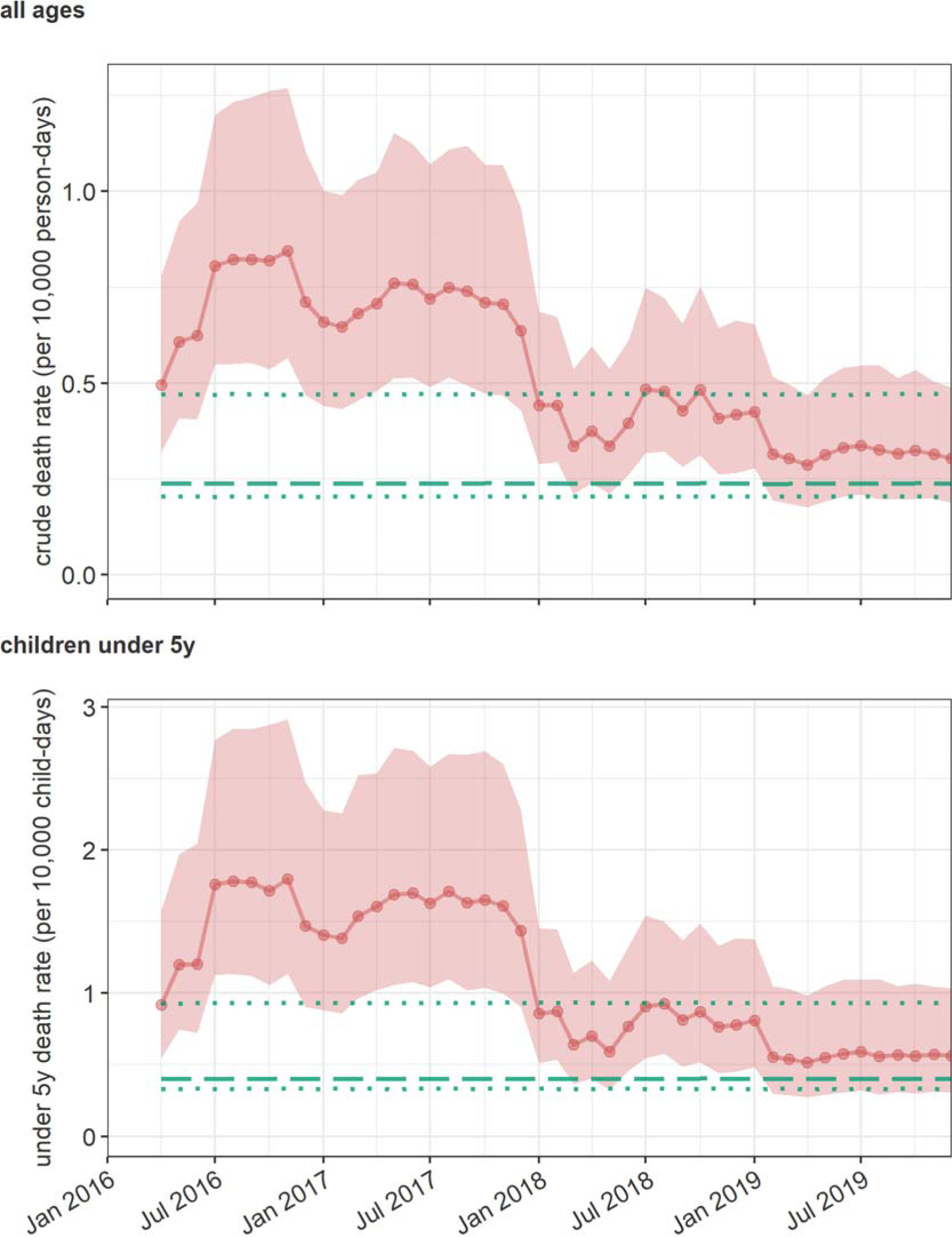
Trends in the estimated crude and under 5y death rates. Red dotted lines and shaded areas indicate the point estimate and 95%CI. The green dashed line is the most likely counterfactual level, and green dotted lines indicate the reasonable best- and worst-case counterfactual scenarios.

Geographically, the highest, but also lowest, CDRs were estimated for LGAs in Borno state (Figure 3). Notably, we projected consistently elevated CDR (compared to the counterfactual or, indeed, national estimates) across Adamawa and Yobe states. Note however that at the LGA level estimates are subject to considerable imprecision; furthermore, chronological trends are obfuscated by these period averages.

**Figure 3.**
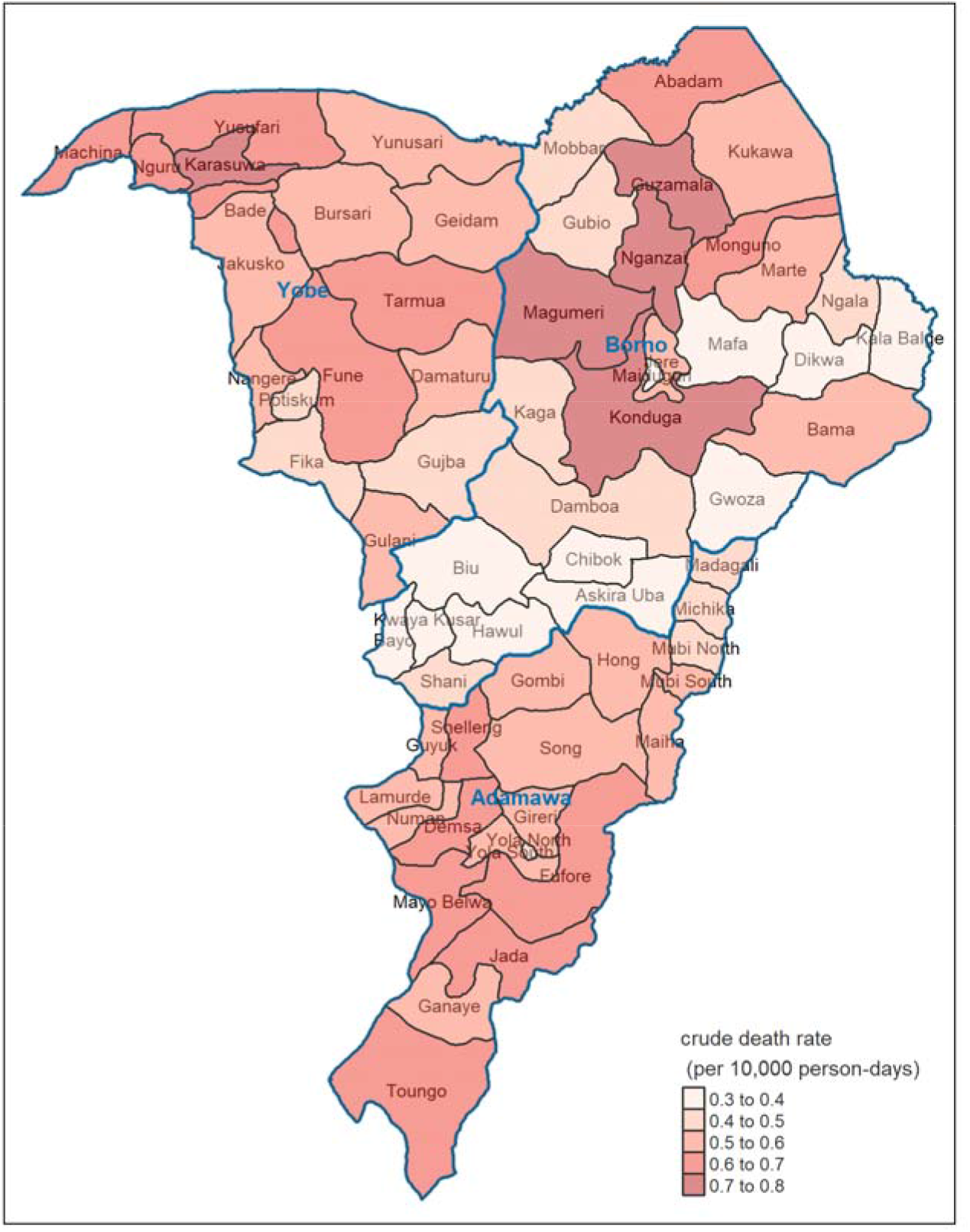
Estimated average CDR during the analysis period, by LGA.

## Discussion

Our analysis finds that a crisis fuelled by armed conflict and further complicated by food insecurity likely caused a very large death toll over four years in Adamawa, Borno and Yobe states, with about half of the excess mortality among children under 5y old. Our central estimate suggests that between 2016 and 2019, the region lost about 4-5% of its population because of the crisis. Death rates were highest in 2016-2017 and reduced to less acute levels by 2019. Despite armed conflict and displacement being focalised in Borno state, we project equally or higher death tolls in Adamawa and Yobe (see Limitations). Generally, estimates are subject to considerable uncertainty, particularly as the counterfactual assumptions are varied.

Over the past decades, few attempts have been made to document mortality at the level of entire crises. However, studies have consistently illustrated the stark impact of armed conflict, displacement and/or food insecurity on human survival. In the Democratic Republic of Congo, successive nationwide surveys estimated CDRs 1.4 to 1.7 times higher than pre-war, with >4M excess deaths between 1998 and 2006 [28, 29]. Surveys in Iraq between 2004 and 2011 found death tolls in the hundreds of thousands, with CDR about 1.5 to 3 times pre-invasion levels [30]. During a famine in Somalia (2010-2012), CDR increased about 5- to 8-fold, with some 260,000 excess deaths [31], while in South Sudan we estimated 380,000 excess deaths, about 1.7 times the counterfactual [32]. Our estimate is however temporally and geographically incomplete, as it omits a long period (2009-2015) of conflict between Nigerian authorities and militant groups, in particular the 2013-2015 escalation period during which large waves of displacement occurred and humanitarian assistance was scarce. It also does not cover the experience of refugees and communities in Niger, Chad and Cameroon affected by what has turned into a transnational conflict.

### Limitations

Our statistical model for CDR appears consistent with the observed data and, importantly, features plausible, dose-response associations. The model for U5DR is less accurate, presumably reflecting sparser input data. The overall estimates rest on strong assumptions that any bias in the input data themselves does not have a substantial influence on the model and its predictions. We did sensitivity analyses to explore two key sources of potential bias: (i) inaccurate estimation of IDP numbers and (ii) under-ascertainment of under 5 years deaths. First, we assumed various levels of under- or over-estimation in IDP figures (from 0.5 to 1.5 as a ratio of actual to observed figures) and population estimates (from 0.7 to 1.3), repeating all downstream analysis steps with the resulting alternative population denominators. Results appear highly sensitivity to bias in population figures, but far less to IDP data bias (Figure S4). Second, we assumed that a proportion up to 50% of under 5 years deaths (neonates and infants in particular) were missed during ground survey household interviews: this is a potential limitation of SMART surveys [33], particularly where questionnaires are administered sub-optimally or child deaths are stigmatising, and may underlie the relatively low proportion of infants among all under 5y deaths within the surveys we analysed. Death tolls among children under 5y would approximately double if only half of childhood deaths had been detected (Figure S13).

Our study does not shed light on cause-specific mortality. Although 25 of the ground surveys collected cause-of-death information from next-of-kin, these surveys were clustered in time and space and were thus not considered a sufficient training and validation dataset for statistical models. However, the very large proportion of injury deaths, particularly in Borno and during 2016, combined with the strikingly higher risk of dying among males than among females, are consistent with violence having been a leading direct cause of mortality. The different databases of insecurity we analysed tallied 10,944 (ACLED), 10,540 (Nigeria Security Tracker), 4859 (Global Terrorism Database) and 13,300 (Nigeria Watch) people killed during our same analysis period (note that these totals reflect not just sensitivity of detection, but also database-specific inclusion criteria). Unexpectedly, none of these sources of data, despite well-documented and systematic procedures for data collection and curation, were correlated with CDR or U5DR in models, in contrast with previous analyses in Somalia [31, 32] and South Sudan [32], where insecurity events were a key overall mortality predictor, and global geospatial associations of armed conflict intensity with child mortality [34]. One explanation for this lack of association could be that insecurity monitoring projects face a common challenge of ascertaining events and deaths in inaccessible areas, and generally where overall mortality is highest; this seems unlikely, as in fact all four sources do show much higher incidence of killings in partially or completely inaccessible LGAs, albeit with somewhat discordant patterns (Supplementary Material). As shown in Figure 5, while the rate of killings during the recall period of ground surveys does somewhat correlate with the proportion of deaths reported as injury-related, there is no visible correlation of either injury-related proportional mortality or the rate of killings with CDR itself. We speculate that the restriction of surveys’ effective sampling frame to accessible, relatively secure areas of LGAs may provide an explanation.

**Figure 5.**
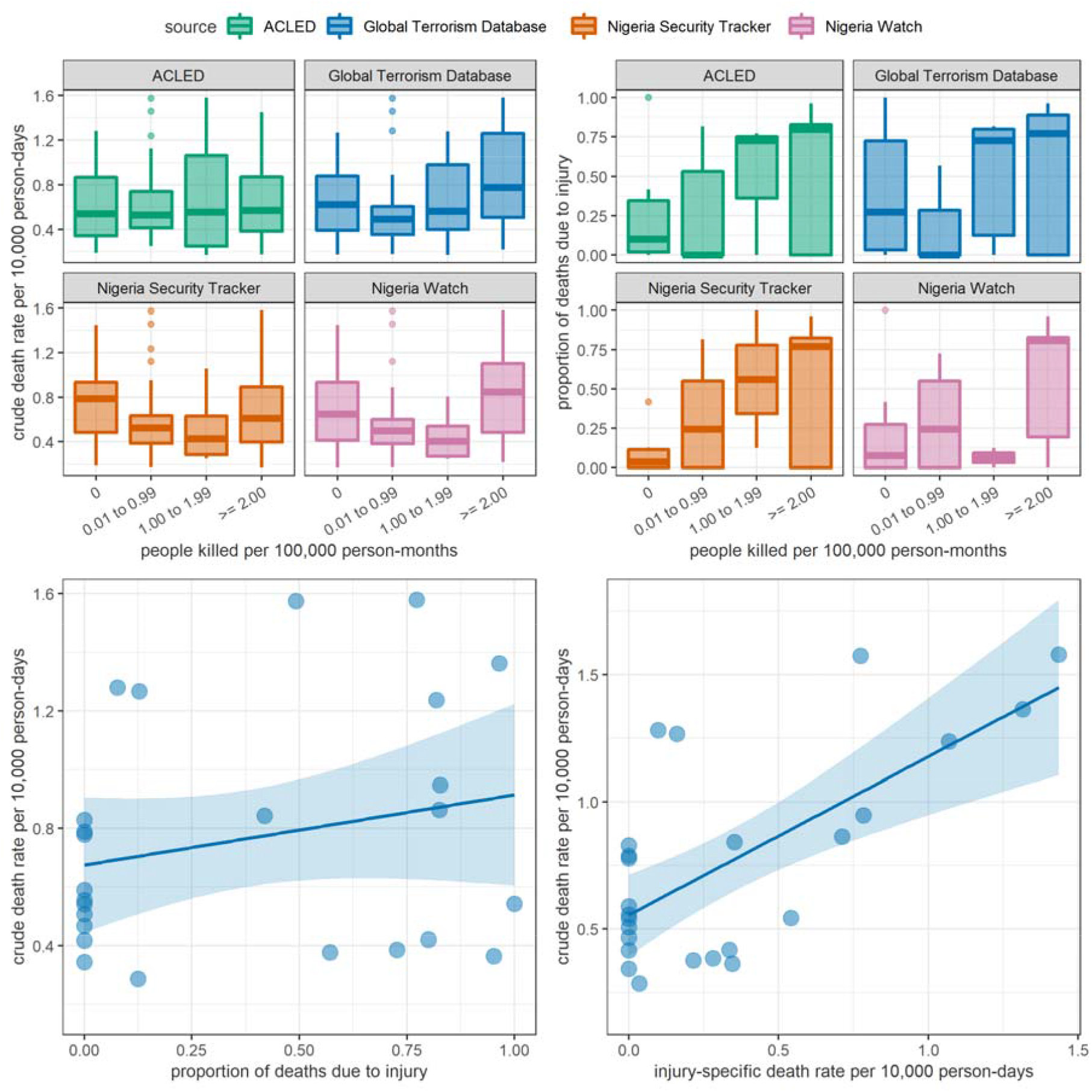
Correlations among different mortality indicators as estimated by ground SMART surveys and the rate of people killed as reported by alternative insecurity monitoring projects. In the top panels, box plots indicate the median and inter-quartile range; whiskers indicate the 95% percentiles. In the bottom panels, straight lines and shaded areas indicate linear regression best fits and 95% confidence intervals, as a crude representation of the extent of correlation. ACLED = Armed Conflict Location & Event Data Project.

The above in turn raises the possibility of bias in our estimates due to inaccessible areas being systematically excluded from ground surveys and measurement of predictor data. We explored this in sensitivity analysis by assuming that in partially or completely inaccessible LGA-months the true CDR and U5DR were up to twice our estimates. Estimates were most sensitive to potential underestimation in Borno state, where access was most constrained (Supplementary Material); overall, the median estimate of excess death toll increased from 490,000 to 800,000 in an extreme bias scenario (doubling of CDR). We consider it plausible that, at least for Borno state, our study does substantially under-estimate mortality.

Despite the very sizeable internal displacement during the analysis period, we also did not find that displacement indicators (proportion of IDPs, arrival or departure rate) were associated with mortality, though globally IDPs consistently have higher death rates than resident populations [27]. Locations with very large IDP concentrations would have been relatively more secure (e.g. Maiduguri and Jere LGAs), and benefited disproportionately from humanitarian assistance or the generosity of local civil society; moreover, during this crisis IDPs have preferentially settled among resident populations or in small camps, avoiding some of the public health threats of large, unplanned camps with no nearby essential services.

More fundamentally, our method requires deliberate definitions of what a crisis consists of, and of counterfactual conditions in its absence. These conditions are in turn set by assuming values of model predictors (and of demographic data) in the absence of the crisis. Specifically, we considered hyperinflation among the conditions contributing to the crisis: indeed, increasing price of cereal, as a proxy of this condition, strongly predicted mortality. As shown in the Supplementary Material, the period of high prices in 2016-2017 was a nationwide phenomenon, and thus arguably not a specific feature of the north-east Nigeria crisis. However, for our central counterfactual scenario we reasoned that in the crisis-affected region food insecurity would have had a disproportionate impact on risk of death given the loss of coping mechanisms, low access to public health services and displacement conditions not encountered by other Nigerian populations. Our central mortality estimates thus reflect the added effect of food price increases; our reasonable-best scenario estimate, by contrast, assumes moderately high food prices as part of the expected baseline.

Lastly, our study was conducted from Europe, with limited field presence and insight into the north-east Nigerian context. It would have been more appropriate to more closely involve Nigerian academic or civil society partners in its conduct and interpretation. For example, this lack of insight could have blinded us to validity problems with the datasets we relied on.

### Conclusions

Estimates of the direct and indirect mortality impact of armed conflict and other crisis conditions are needed to appraise the appropriateness of humanitarian responses, but also for governments, combatants and societies to appreciate the consequences of these crises and of military tactics used to prosecute wars. More fundamentally, they help to memorialise those who died and construct objective historical accounts. Our attempt at deriving such an estimate for north-east Nigeria should be critiqued and interpreted in light of its limitations. We contend, however, that generating evidence on the human toll of this very large crisis warrants the involvement of several independent groups, perhaps leveraging different methods. Generally, mortality estimation should become a predictable, systematic public health information service across all crisis settings. This requires concerted work by scientific groups and sustained, long-term funding support.

## Data Availability

Statistical code and datasets to implement the analysis using R software are available online on: these allow users to vary various data management and modelling parameters by modifying their values on Microsoft Excel input files.

https://github.com/francescochecchi/mortality_small_area_estimation/tree/nga

## Additional information

## Acknowledgments

We are grateful to Anna Carnegie for project management support, and to the Nigerian government, United Nations, Action Against Hunger and other organisations who shared datasets or facilitated access to these.

## Disclaimer

Geographical names and boundaries presented in this report are used solely for the purpose of producing scientific estimates, and do not necessarily represent the views or official positions of the authors, the London School of Hygiene and Tropical Medicine, any of the agencies that have supplied data for this analysis, or the donor. The authors are solely responsible for the analyses presented here, and acknowledgment of data sources does not imply that the agencies or individuals providing data endorse the results of the analysis.

## References

1. Onuoha FC. Why Do Youth Join Boko Haram? US Institute of Peace; 2014.

2. Kwaja C. Nigeria’s Pernicious Drivers of Ethno-Religious Conflict. Washington, DC: Africa Center for Strategic Studies; 2011.

3. Obe A. Environmental Degradation, Climate Change and Conflict: The Lake Chad Basin Area. International Crisis Group; 2016.

4. Nigeria: 2020 Humanitarian Needs Overview. Abuja: United Nations Office for Coordination of Humanitarian Affairs; 2019.

5. Kurtzer J. OUT OF SIGHT. Center for Strategic and International Studies (CSIS); 2020.

6. McIlreavy P, Schopp J. A collective shame: the response to the humanitarian crisis in north-eastern Nigeria. Humanitarian Practice Network.

7. Stoddard A, Harvey P, Czwarno M, Breckenridge M. Humanitarian access SCORE report: northeast Nigeria. Survey on the coverage, operational reach, and effectiveness of humanitarian aid. Humanitarian Outcomes; 2020.

8. Food insecurity reaches extreme level in pockets of Nigeria’s Borno State. Washington, DC: Famine Early Warning Systems Network; 2016.

9. Cadre Harmonisé Update Analysis in Adamawa, Borno and Yobe States of Nigeria. Abuja: Food and Agriculture Organization; 2016.

10. Special alert on Borno state, Nigeria: Urgent humanitarian action needed to respond to an elevated risk of famine. Integrated Food Security Phase Classification; 2016.

11. Omole O, Welye H, Abimbola S. Boko Haram insurgency: implications for public health. The Lancet. 2015;385:941.

12. Checchi F, Warsame A, Treacy-Wong V, Polonsky J, van Ommeren M, Prudhon C. Public health information in crisis-affected populations: a review of methods and their use for advocacy and action. Lancet. 2017;390:2297–313.

13. Checchi F. Estimation of population mortality in crisis-affected populations: Guidance for humanitarian coordination mechanisms. Geneva: World Health Organization; 2018.

14. Rao JNK, Molina I. Small Area Estimation: Rao/Small Area Estimation. Hoboken, NJ, USA: John Wiley & Sons, Inc; 2015.

15. Checchi F, Testa A, Gimma A, Koum-Besson E, Warsame A. A method for small-area estimation of population mortality in settings affected by crises. Popul Health Metrics. 2022;20:4.

16. R Core Team. R: A Language and Environment for Statistical Computing. 2020.

17. Standardised Monitoring and Assessment of Relief and Transitions (SMART). Measuring Mortality, Nutritional Status, and Food Security in Crisis Situations: SMART Methodology. https://smartmethodology.org/. Accessed 14 Feb 2021.

18. Cairns KL, Woodruff BA, Myatt M, Bartlett L, Goldberg H, Roberts L. Cross-sectional survey methods to assess retrospectively mortality in humanitarian emergencies. Disasters. 2009;33:503–21.

19. Funk C, Peterson P, Landsfeld M, Pedreros D, Verdin J, Shukla S, et al. The climate hazards infrared precipitation with stations—a new environmental record for monitoring extremes. Scientific Data. 2015;2:150066.

20. Raleigh C, Linke A, Hegre H, Karlsen J. Introducing ACLED: An Armed Conflict Location and Event Dataset: Special Data Feature. Journal of Peace Research. 2010;47:651–60.

21. Touray K, Mkanda P, Tegegn SG, Nsubuga P, Erbeto TB, Banda R, et al. Tracking Vaccination Teams During Polio Campaigns in Northern Nigeria by Use of Geographic Information System Technology: 2013-2015. J Infect Dis. 2016;213 Suppl 3 Suppl 3:S67–72.

22. Higgins J, Adamu U, Adewara K, Aladeshawe A, Aregay A, Barau I, et al. Finding inhabited settlements and tracking vaccination progress: the application of satellite imagery analysis to guide the immunization response to confirmation of previously-undetected, ongoing endemic wild poliovirus transmission in Borno State, Nigeria. Int J Health Geogr. 2019;18:11–11.

23. Barau I, Zubairu M, Mwanza MN, Seaman VY. Improving Polio Vaccination Coverage in Nigeria Through the Use of Geographic Information System Technology. The Journal of Infectious Diseases. 2014;210 suppl_1:S102–10.

24. Gneiting T, Raftery AE. Strictly Proper Scoring Rules, Prediction, and Estimation. Journal of the American Statistical Association. 2007;102:359–78.

25. Brooks ME, Kristensen K, Benthem KJ van, Magnusson A, Berg CW, Nielsen A, et al. glmmTMB Balances Speed and Flexibility Among Packages for Zero-inflated Generalized Linear Mixed Modeling. The R Journal. 2017;9:378–400.

26. United Nations D of E, Social Affairs PD. World Population Prospects - Population Division - United Nations. 2019.

27. Heudtlass P, Speybroeck N, Guha-Sapir D. Excess mortality in refugees, internally displaced persons and resident populations in complex humanitarian emergencies (1998-2012) - insights from operational data. Confl Health. 2016;10:15.

28. Coghlan B, Ngoy P, Mulumba F, Hardy C, Bemo VN, Stewart T, et al. Update on mortality in the Democratic Republic of Congo: results from a third nationwide survey. Disaster Med Public Health Prep. 2009;3:88–96.

29. Coghlan B, Brennan RJ, Ngoy P, Dofara D, Otto B, Clements M, et al. Mortality in the Democratic Republic of Congo: a nationwide survey. The Lancet. 2006;367:44–51.

30. Hagopian A, Flaxman AD, Takaro TK, Esa Al Shatari SA, Rajaratnam J, Becker S, et al. Mortality in Iraq associated with the 2003-2011 war and occupation: findings from a national cluster sample survey by the university collaborative Iraq Mortality Study. PLoS Med. 2013;10:e1001533.

31. Checchi F, Robinson, Courtland. Mortality among populations of southern and central Somalia affected by severe food insecurity and famine during 2010-2012 - Somalia. Nairobi: Food and Agriculture Organization; 2013.

32. Checchi F, Testa, Adrienne, Warsame, Abdihamid, Quach, Le, Burns, Rachel. Estimates of crisis-attributable mortality in South Sudan, December 2013-April 2018: A statistical analysis - South Sudan. ReliefWeb. 2018. https://reliefweb.int/report/south-sudan/estimates-crisis-attributable-mortality-south-sudan-december-2013-april-2018. Accessed 11 Jan 2021.

33. Working Group for Mortality Estimation in Emergencies. Wanted: studies on mortality estimation methods for humanitarian emergencies, suggestions for future research. Emerg Themes Epidemiol. 2007;4:9.

34. Wagner Z, Heft-Neal S, Bhutta ZA, Black RE, Burke M, Bendavid E. Armed conflict and child mortality in Africa: a geospatial analysis. The Lancet. 2018;392:857–65.

