## Supplementary Material for "The death toll of armed conflict and food insecurity in north-east Nigeria, 2016-2019: a statistical study"

### Methods

#### Geographic strata

Table S1 lists the different geographic units associated with each LGA, including state, domain (used as the sampling universe of most mortality surveys) and nearest market.

Table S1. List of geographic units and equivalencies.

| state | domain | subdomain | LGA | market | criterion for assigning the LGA to the market |
| --- | --- | --- | --- | --- | --- |
| Adamawa | Southern Adamawa | Southern Adamawa | **Demsa** | Mubi | Closest market to the LGA |
| Adamawa | Southern Adamawa | Southern Adamawa | **Fufore** | Mubi | Closest market to the LGA |
| Adamawa | Southern Adamawa | Southern Adamawa | **Ganaye** | Mubi | Closest market to the LGA |
| Adamawa | Northern Adamawa | Northern Adamawa B | **Gireri** | Mubi | Same domain as the market |
| Adamawa | Southern Adamawa | Southern Adamawa | **Gombi** | Mubi | Closest market to the LGA |
| Adamawa | Southern Adamawa | Southern Adamawa | **Guyuk** | Mubi | Closest market to the LGA |
| Adamawa | Northern Adamawa | Northern Adamawa B | **Hong** | Mubi | Same domain as the market |
| Adamawa | Southern Adamawa | Southern Adamawa | **Jada** | Mubi | Closest market to the LGA |
| Adamawa | Southern Adamawa | Southern Adamawa | **Lamurde** | Mubi | Closest market to the LGA |
| Adamawa | Northern Adamawa | Northern Adamawa A | **Madagali** | Mubi | Same domain as the market |
| Adamawa | Northern Adamawa | Northern Adamawa A | **Maiha** | Mubi | Same domain as the market |
| Adamawa | Southern Adamawa | Southern Adamawa | **Mayo Belwa** | Mubi | Closest market to the LGA |
| Adamawa | Northern Adamawa | Northern Adamawa A | **Michika** | Mubi | Same domain as the market |
| Adamawa | Northern Adamawa | Northern Adamawa A | **Mubi North** | Mubi | Same domain as the market |
| Adamawa | Northern Adamawa | Northern Adamawa B | **Mubi South** | Mubi | Same domain as the market |
| Adamawa | Southern Adamawa | Southern Adamawa | **Numan** | Mubi | Closest market to the LGA |
| Adamawa | Southern Adamawa | Southern Adamawa | **Shelleng** | Mubi | Closest market to the LGA |
| Adamawa | Northern Adamawa | Northern Adamawa B | **Song** | Mubi | Same domain as the market |
| Adamawa | Southern Adamawa | Southern Adamawa | **Toungo** | Mubi | Closest market to the LGA |
| Adamawa | Southern Adamawa | Southern Adamawa | **Yola North** | Mubi | Closest market to the LGA |
| Adamawa | Southern Adamawa | Southern Adamawa | **Yola South** | Mubi | Closest market to the LGA |
| Borno | Northern Borno | Northern Borno | **Abadam** | Maiduguri | Closest market to the LGA |
| Borno | Southern Borno | Southern Borno | **Askira Uba** | Biu | Same domain as the market |
| Borno | Eastern Borno | Eastern Borno | **Bama** | Maiduguri | Closest market to the LGA |
| Borno | Southern Borno | Southern Borno | **Bayo** | Biu | Same domain as the market |
| Borno | Southern Borno | Southern Borno | **Biu** | Biu | Same domain as the market |
| Borno | Southern Borno | Southern Borno | **Chibok** | Biu | Same domain as the market |
| Borno | Central Borno | Central Borno A | **Damboa** | Maiduguri | Closest market to the LGA |
| Borno | Eastern Borno | Eastern Borno | **Dikwa** | Maiduguri | Closest market to the LGA |
| Borno | Central Borno | Central Borno B | **Gubio** | Maiduguri | Closest market to the LGA |
| Borno | Northern Borno | Northern Borno | **Guzamala** | Maiduguri | Closest market to the LGA |
| Borno | Eastern Borno | Eastern Borno | **Gwoza** | Maiduguri | Closest market to the LGA |
| Borno | Southern Borno | Southern Borno | **Hawul** | Biu | Same domain as the market |
| Borno | MMC/Jere | MMC/Jere | **Jere** | Maiduguri | Same domain as the market |
| Borno | Central Borno | Central Borno A | **Kaga** | Maiduguri | Closest market to the LGA |
| Borno | Eastern Borno | Eastern Borno | **Kala Balge** | Maiduguri | Closest market to the LGA |
| Borno | Central Borno | Central Borno A | **Konduga** | Maiduguri | Closest market to the LGA |
| Borno | Northern Borno | Northern Borno | **Kukawa** | Maiduguri | Closest market to the LGA |
| Borno | Southern Borno | Southern Borno | **Kwaya Kusar** | Biu | Same domain as the market |
| Borno | Central Borno | Central Borno B | **Mafa** | Maiduguri | Closest market to the LGA |
| Borno | Central Borno | Central Borno B | **Magumeri** | Maiduguri | Closest market to the LGA |
| Borno | MMC/Jere | MMC/Jere | **Maiduguri** | Maiduguri | Same domain as the market |
| Borno | Central Borno | Central Borno B | **Marte** | Maiduguri | Closest market to the LGA |
| Borno | Northern Borno | Northern Borno | **Mobbar** | Maiduguri | Closest market to the LGA |
| Borno | Central Borno | Central Borno B | **Monguno** | Maiduguri | Closest market to the LGA |
| Borno | Eastern Borno | Eastern Borno | **Ngala** | Maiduguri | Closest market to the LGA |
| Borno | Northern Borno | Northern Borno | **Nganzai** | Maiduguri | Closest market to the LGA |
| Borno | Southern Borno | Southern Borno | **Shani** | Biu | Same domain as the market |
| Yobe | Central Yobe | Central Yobe | **Bade** | Potiskum | Closest market to the LGA |
| Yobe | Central Yobe | Central Yobe | **Bursari** | Damaturu | Closest market to the LGA |
| Yobe | Southern Yobe | Southern Yobe A | **Damaturu** | Damaturu | Closest market to the LGA |
| Yobe | Southern Yobe | Southern Yobe A | **Fika** | Potiskum | Closest market to the LGA |
| Yobe | Southern Yobe | Southern Yobe A | **Fune** | Potiskum | Closest market to the LGA |
| Yobe | Central Yobe | Central Yobe | **Geidam** | Damaturu | Closest market to the LGA |
| Yobe | Southern Yobe | Southern Yobe B | **Gujba** | Damaturu | Closest market to the LGA |
| Yobe | Southern Yobe | Southern Yobe B | **Gulani** | Potiskum | Closest market to the LGA |
| Yobe | Central Yobe | Central Yobe | **Jakusko** | Potiskum | Closest market to the LGA |
| Yobe | Northern Yobe | Northern Yobe A | **Karasuwa** | Potiskum | Closest market to the LGA |
| Yobe | Northern Yobe | Northern Yobe B | **Machina** | Potiskum | Closest market to the LGA |
| Yobe | Southern Yobe | Southern Yobe A | **Nangere** | Potiskum | Closest market to the LGA |
| Yobe | Northern Yobe | Northern Yobe A | **Nguru** | Potiskum | Closest market to the LGA |
| Yobe | Southern Yobe | Southern Yobe A | **Potiskum** | Potiskum | Closest market to the LGA |
| Yobe | Southern Yobe | Southern Yobe A | **Tarmua** | Damaturu | Closest market to the LGA |
| Yobe | Northern Yobe | Northern Yobe B | **Yunusari** | Potiskum | Closest market to the LGA |
| Yobe | Northern Yobe | Northern Yobe B | **Yusufari** | Potiskum | Closest market to the LGA |

#### SMART mortality surveys

Figure S1 shows the LGA-month coverage of the 70 SMART mortality surveys included in the analysis. No surveys could be acquired in 2019 due to the time-limited extent of approvals to access data. We otherwise believe that, with the exception of a small number of surveys done by Médecins Sans Frontières in IDP settlements, we were able to secure the vast majority of mortality data available during 2016-2018.

We cleaned each dataset to remove obviously wrong entries and re-analysed each survey using a fixed-effects Poisson regression, with standard errors adjusted for intra-cluster correlation as per the R survey package. We computed a survey weight in the range 0 to 1 for each observation, equal to the anthropometric quality score automatically generated by ENA software during validation of each survey (this score considers various patterns in the data suggestive of adequate staff training and fieldwork [1]; we normalised scores to 1; see Figure S2), multiplied by the proportion of the intended sampling universe that was actually reached during the survey, as per survey report (or 1.0 if the entire sampling universe was reached).

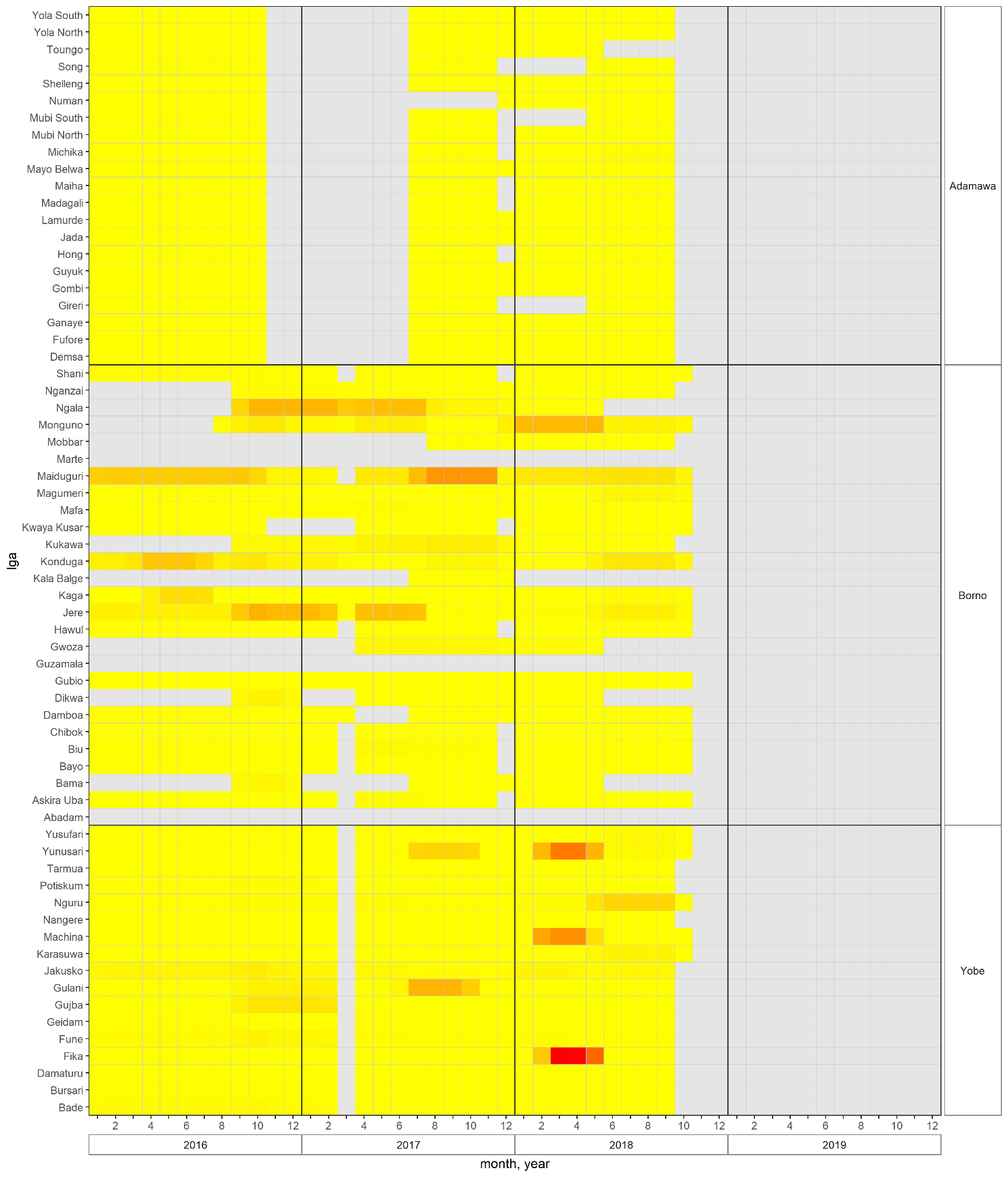

Figure S1. Coverage of SMART mortality surveys, by state and LGA. Heat colours denote months falling within the recall period of one or more surveys, with increasing colour intensity proportional to the precision of estimates.

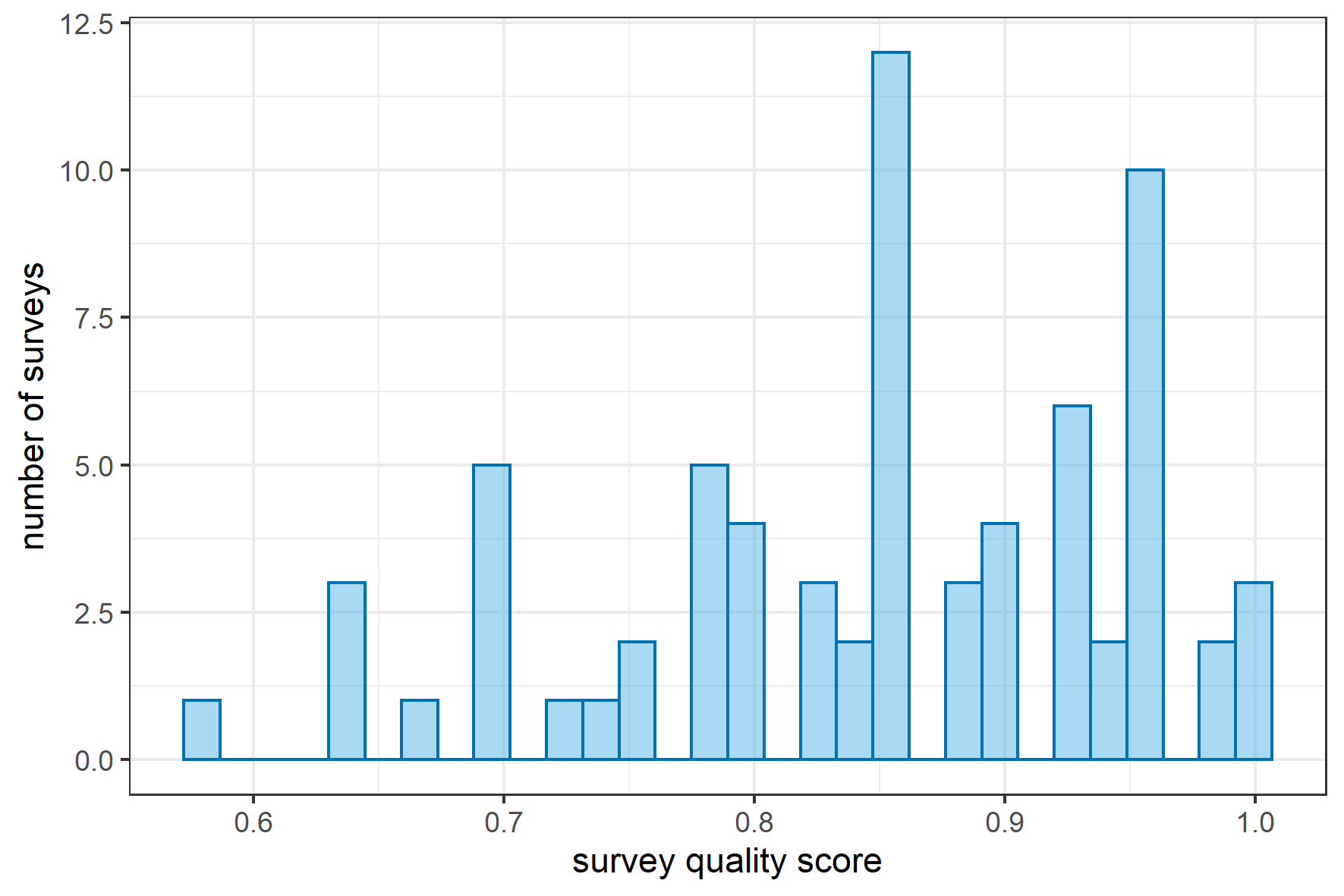

Figure S2. Distribution of quality score of SMART mortality surveys included in the analysis.

#### Reconstruction of population denominators

Table S2 lists the different sources of information we drew on to estimate the number of people present in any LGA during any given month, as well as the number of IDPs and refugees leaving, arriving (or returning) into and currently displaced within each LGA (note that internal displacement could also be within the LGA itself).

Table S2. Sources of data on population and displacement.

| Source (URL) | Reference date | Details |
| --- | --- | --- |
| Population estimates | | |
| National Bureau of Statistics  (<https://data.humdata.org/dataset/nigeria-2016-population-data>) | June 2016 | Projections of the 2006 national census, supported by humanitarian actors. |
| Geo-Referenced Infrastructure and Demographic Data for Development (GRID^3^)  (<https://grid3.gov.ng/>) | August 2019 | The method combines census data, ad hoc micro-census surveys, ground settlement data and remote sensing information into a statistical model, partly informed by the AfriPop approach, that generates population estimates for 100m^2^ grids. It builds on previous estimation work in northern Nigeria [2]. |
| Facebook Data for Good (<https://data.humdata.org/dataset/highresolutionpopulationdensitymaps-nga> ) | November 2016 | Population estimates for 30m2 grids, based on remote sensing approaches combined with available census and demographic survey estimates. The method is based on neural networks and rests on enumerating inhabited structures. It is explicitly conceived for use by humanitarian actors [3]. |
| Emergency Operations Centre, northeast Nigeria | November 2018 | Using polio vaccination teams as the backbone, these estimates are derived by tracking settlement habitation through a combination of remote sensing and field data [4]. |
| IDP movements | | |
| International Organisation for Migration Displacement Tracking Matrix (<https://dtm.iom.int/>) | February 2015 to October 2019 | Regular (mostly every two months) assessments of all known and accessible IDP settlements, collecting data on reported numbers of IDPs, their year of arrival and main LGA of origin. |
| Refugee movements | | |
| United Nations High Commissioner for Refugees | January 2015 to December 2019 | Approximated by the authors based on document review and triangulation of different datasets, after a systematic review of UNHCR Nigeria crisis documents on the [www.reliefweb.org](http://www.reliefweb.org) humanitarian document repository. |

Briefly, for each LGA we forward- or back-calculated population from the time point at which each of the alternative population estimate sources was centred, using the following equation:

$$N_{t+1}=N_{t}(1+g)+I_{A, t}-I_{D, t}+R_{A, t}-R_{D, t}$$

where $N$ is total population, $g$ the assumed growth rate, $I$ internally displaced persons and $R$ refugees ($A$ = arriving from another LGA, $D$ = departing to another LGA). We then took a weighted average of the four time series arising from each population source, with the weight corresponding to a quality score comprised of various criteria [5]. Lastly, we computed the population of children under 5y using the mean proportion in this age group (20%) reported by the SMART surveys we analysed.

In order to estimate $I$ flows, we first aggregated IDP groups within each LGA by their year of arrival and LGA of origin, as per the DTM database. Setting 1 Jan 2015 as the start date for population reconstruction, we assumed that everyone who had arrived before then was present at the start date; similarly, further arrivals during 2015 were present as of Jan 2016; and so on. We then interpolated prevalent IDP data as reported by each successive round of assessment, so as to construct monthly time series of IDPs by LGA of arrival and LGA of origin, which we used to compute flows from each LGA to other LGAs for each month. We then used these flow time series in the above equation.

As a final adjustment, we applied a correction to IDP time series whenever reconstructed population for any LGA and any alternative population source became negative, presumably because of systematic error in IDP counting (this occurred in 12/65 LGAs). The correction downward-scaled the time series so as to achieve a minimum of $N$ = 0 for all population sources and LGAs. More detail is available on request.

Figure S3 shows trends in the estimated population by LGA, showing that the least stable patterns occurred in Borno state, where large waves of displacement and return occurred during the period. Figure S4 shows, at state level, the differences in estimates derived from each the four population sources: notably, these differences were not consistent across the three states. Lastly, Figure S5 shows the percentage of each state’s population that consisted of IDPs, indicating a relatively stable pattern over the period. We separately estimate that there were 138,000 refugees from Adamawa, Borno and Yobe in neighbouring countries (Niger, Chad, Cameroon) as of January 2015. Refugee numbers had risen progressively to 239,000 by December 2019.

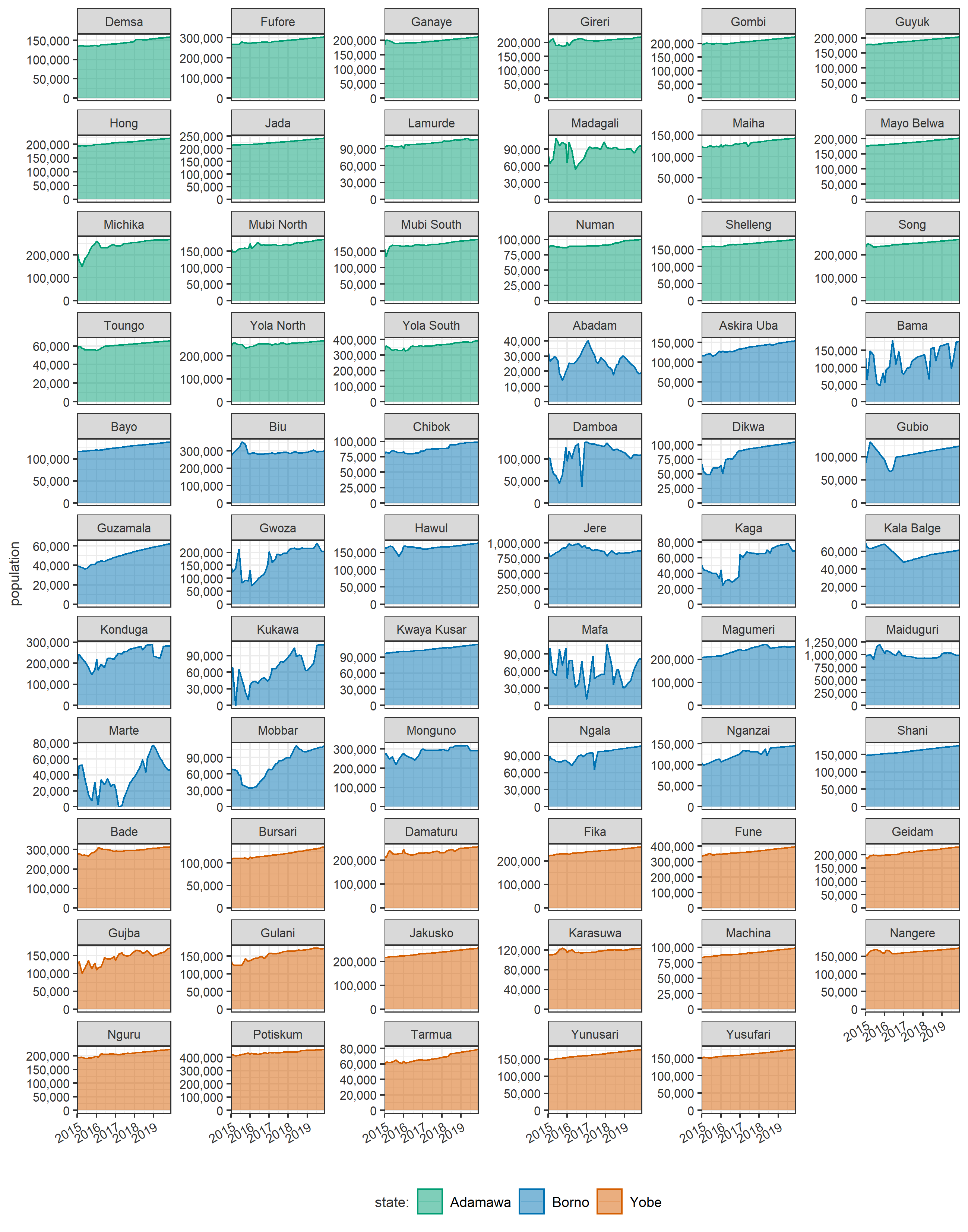

Figure S3. Evolution of estimated population, by LGA.

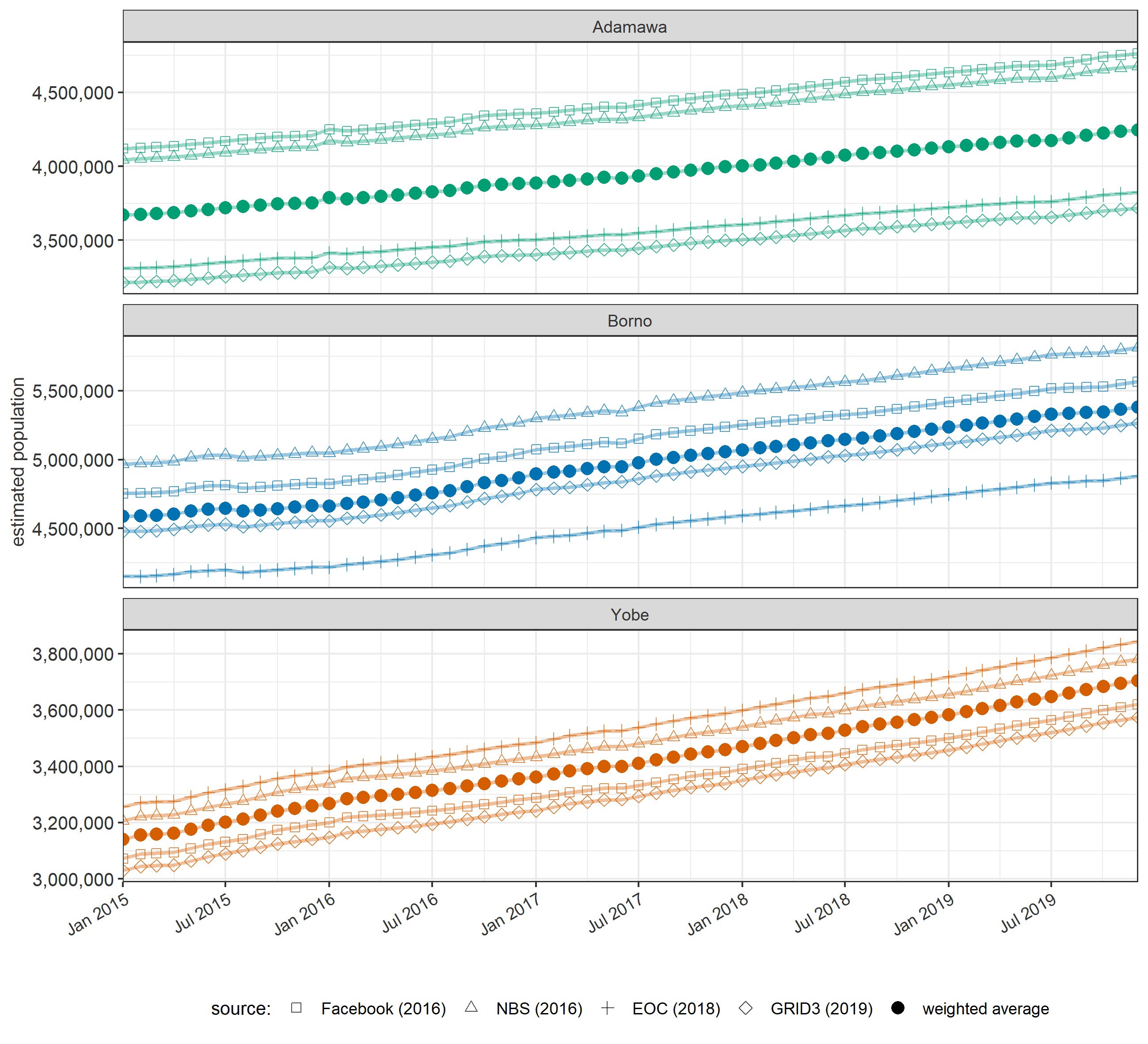

Figure S4. Evolution of estimated population, by alternative population source and state.

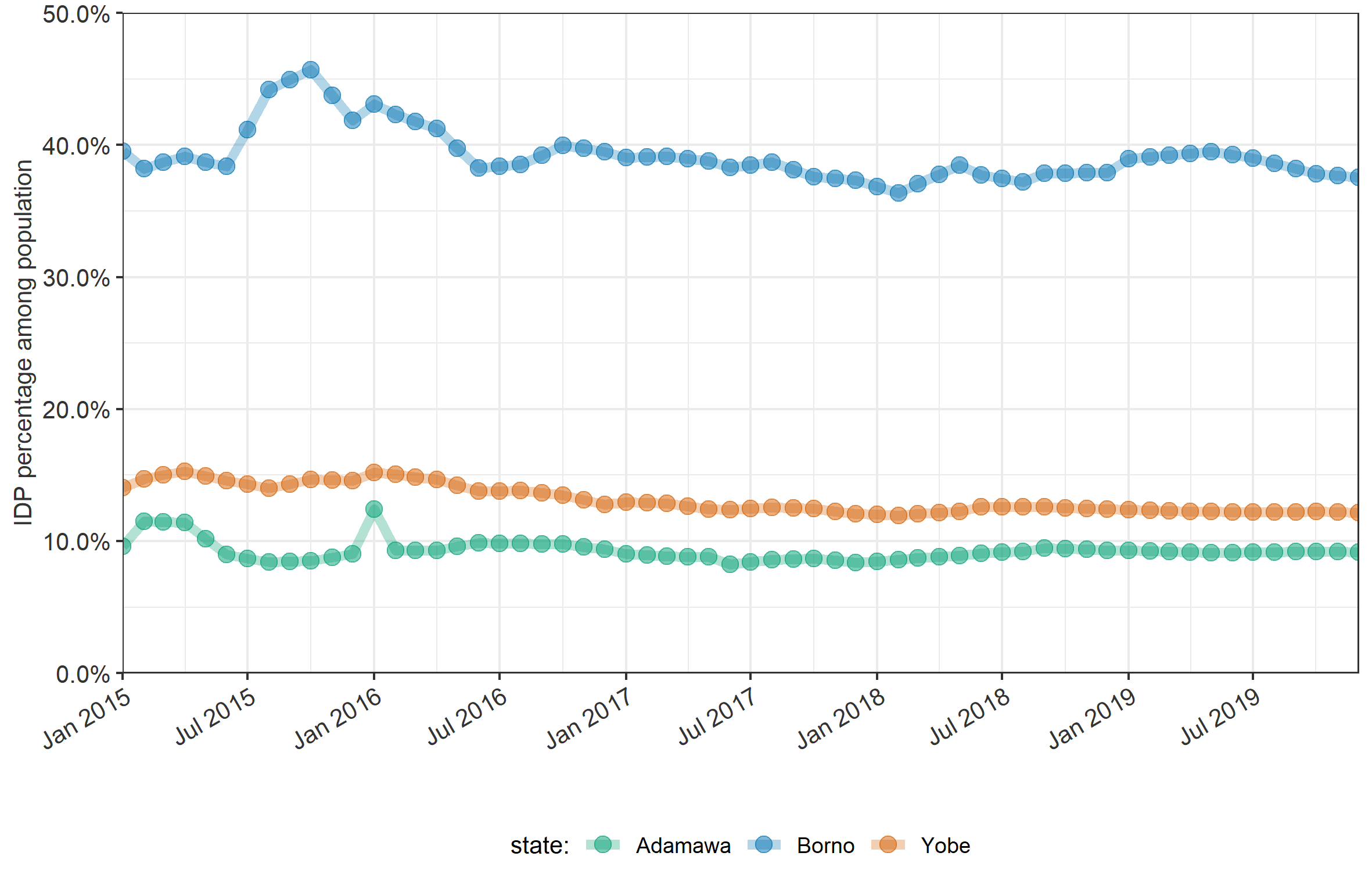

Figure S5. Evolution of the estimated proportion of the population that are IDPs, by state.

#### Completeness of predictor datasets

Figure S6 and Figure S7 show the completeness of candidate predictor datasets, by month (i.e. proportion of LGAs that have complete data for any given month) and LGA (i.e. proportion of months for which a given LGA has a complete data), respectively.

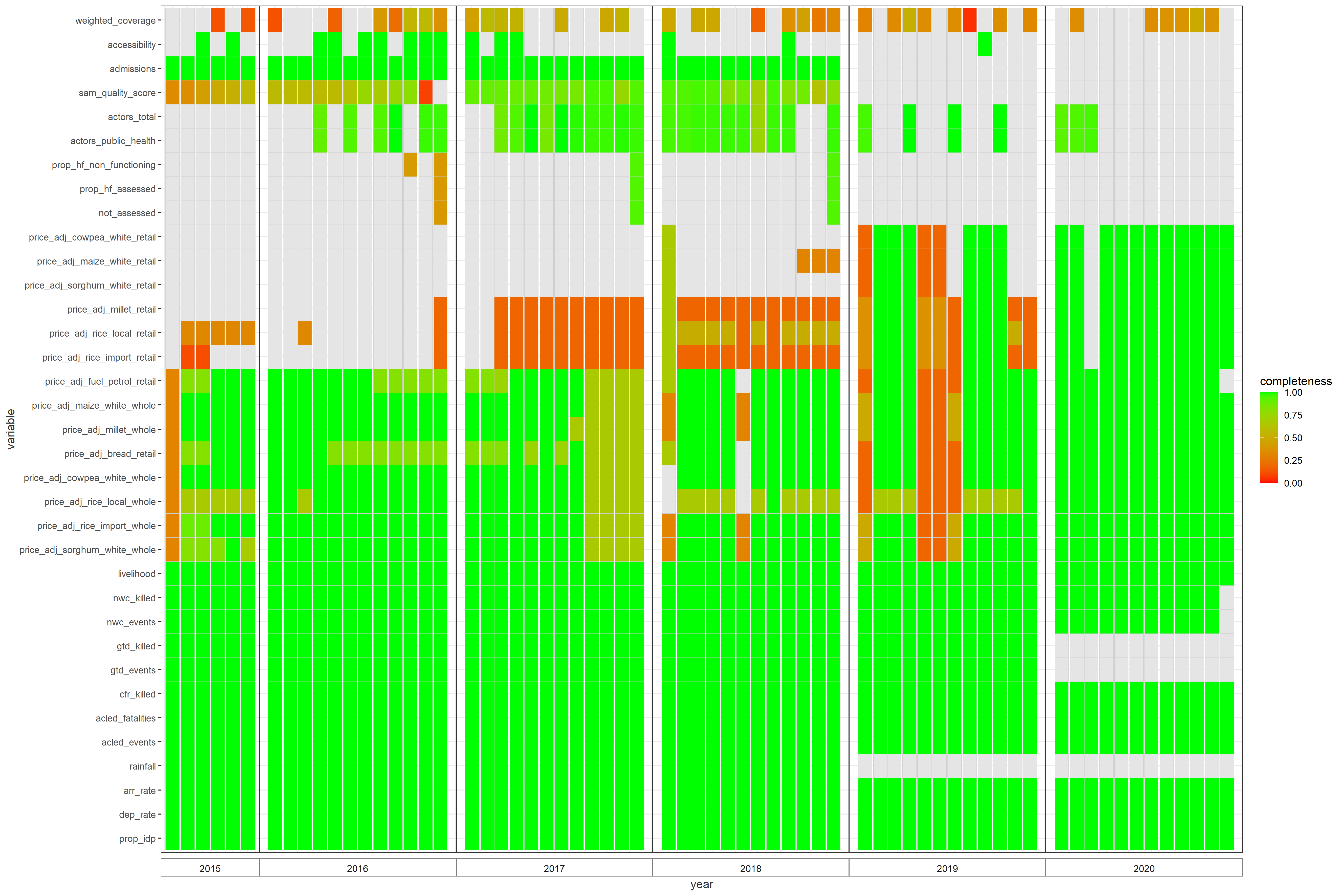

Figure S6. Completeness of candidate predictor datasets, by month, prior to imputation, interpolation or smoothing.

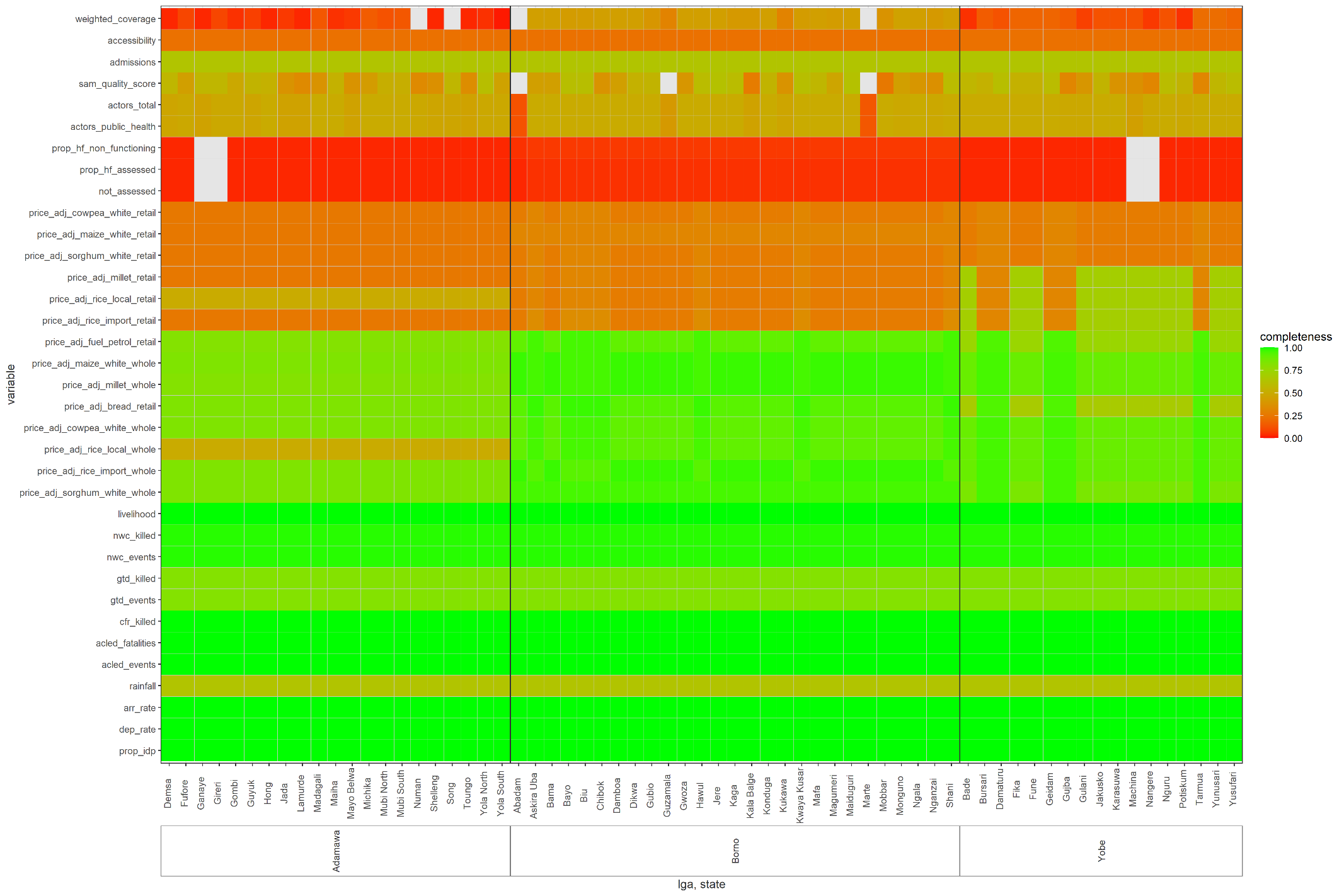

Figure S7. Completeness of candidate predictor datasets, by LGA, prior to imputation, interpolation or smoothing.

#### Management of predictor datasets

##### Market prices

*Data management.* The World Food Programme’s dataset of market prices consists of monthly observations from a purposive selection of markets across Nigeria, with relatively more markets being followed in crisis-affected states. While data are available from 2002, in Adamawa, Borno and Yobe states they are very sparse prior to 2015.

We selected the most frequently reported-on commodities in the dataset: gasoline fuel, bread, white cowpea, white maize, millet, local rice, imported rice and white sorghum. For some of these, both wholesale and retail prices were available.

We computed an inflation-adjusted standardised price for 1Kg of each cereal staple, 1L of fuel and 1 loaf of bread as follows. First, we divided reported cereal prices by their unit to come up with a standardised price per Kg. Second, we converted the price to USD equivalent using historical monthly NGN to USD exchange rates obtained from <https://fxtop.com/en/historical-exchange-rates.php> . Lastly, we adjusted the price for USD inflation (<https://www.in2013dollars.com/us/inflation/2010?amount=1>) to come up with a price in 2010 USD.

While 12 individual markets were featured in the database for Adamawa, Borno and Yobe states, in practice only five had continuous or at least intermittent data coverage from 2015 onwards: Mubi (Adamawa), Biu and Maiduguri (Borno), Damaturu and Potiskum (Yobe). We assigned each LGA to one of these markets based on proximity criteria (Table S1). Each LGA price series was subjected to moderate spline smoothing to correct for possible data entry errors and interpolate between missing observations.

*Setting counterfactual values.* After inspecting the market-specific time series of maize price (Figure S8), we decided for our most likely scenario to take the median values for each market (and thus the LGA attributed to the market) during the low-price periods of January to June 2015 and 2018-2019: this reflects an implicit decision that the price increase should be considered part of the crisis conditions affecting north-east Nigeria. We took the minimum of each market time series as the best-case scenario, and the median of the entire series between 2015-2019 (which therefore includes the price increase period) as the worst-case scenario.

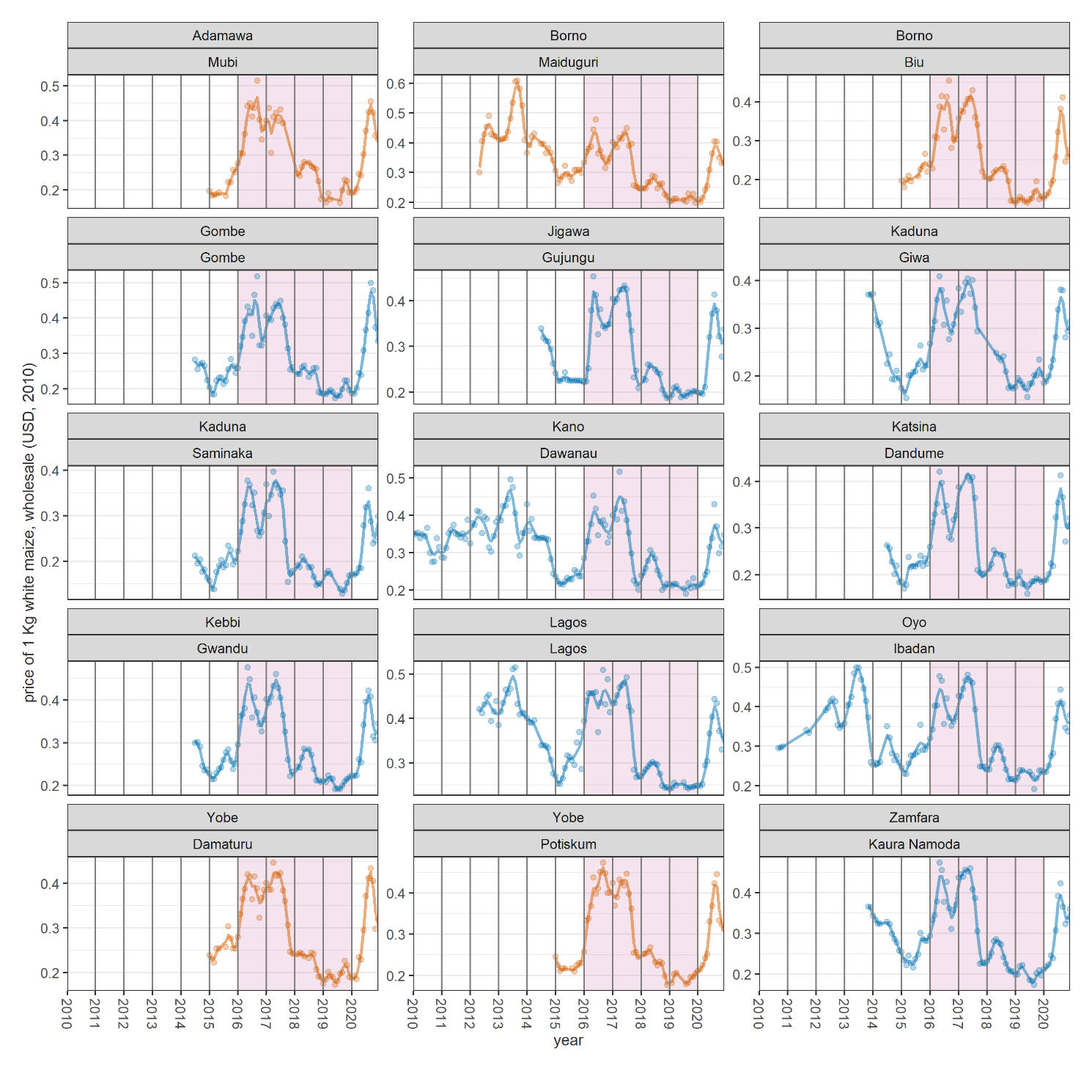

Figure S8. Trends in the price of 1Kg of white maize, at wholesale price (sale unit: 100 Kg), expressed in USD as of 2010, by market. Dots are actual observations and lines are smoothed series. Shading denotes the mortality analysis period.

##### Vaccination coverage

*Data management.* Nigeria’s Vaccination Tracking System (VTS) was developed to help national and state emergency polio operations centres to estimate vaccination coverage and identify areas in need of supplemental vaccination activities. The VTS equips vaccination outreach teams with GPS devices that track the teams’ movements. During their field visits, these teams also collect geographic and demographic information on settlements they visit, which is used not only to estimate vaccination coverage, but also to update population estimates. ‘Vaccination geo-coverage’ during any given time window is quantified as the proportion of 100 m^2^ grids in targeted areas that are visited, based on GPS tracks. The system has more recently been used during seasonal malaria chemoprophylaxis campaigns, and as such more generically produced estimates of the geo-coverage of community-based public health interventions.

We downloaded the ‘time-trend-analysis’ dataset from the now-defunct VTS website (<http://vts.eocng.org/ChronicalCoverage/Index> ). The dataset is organised by ward (sub-unit within LGA and campaign (e.g. August 2017 polio mass vaccination): for each such instance, the dataset reports geo-coverage (estimated as above) and the number of ‘geo-coverage denominators’, roughly equivalent to similar-size population units, and which we used as weights to come up with a weighted geo-coverage by LGA and month. We then did automatic imputation to infer missing values of geo-coverage, which occurred mainly over the intervals between successive campaigns: for this we used the Random Forest method as implemented by the R mice package [6] (<https://amices.org/mice/>), with 5 chains of 20 iterations, and using all available predictors to inform the imputation.

*Setting counterfactual values.* To set counterfactual values, we considered geo-coverage values in states other than Adamawa, Borno and Yobe. After coming up with LGA-month weighted geo-coverage, as above, we retained LGA time series from 2012-2020 that had at least four observations. The resulting dataset contained eligible time series from 49 LGAs in Adamawa, Borno and Yobe and 161 LGAs in 11 other states (Bauchi, Gombe, Jigawa, Kaduna, Kano, Katsina, Kebbi, Kogi, Kwara, Sokoto, Zamfara). We then fitted, for both (i) Adamawa, Borno and Yobe and (ii) all the other states, a generalised additive growth model of geo-coverage (assumed to be normally distributed) as a function of increasing time (month), with time nested within LGA as a random effect, and unity-normalised geo-coverage denominator weights. Model-predicted geo-coverage is shown in Figure S9.

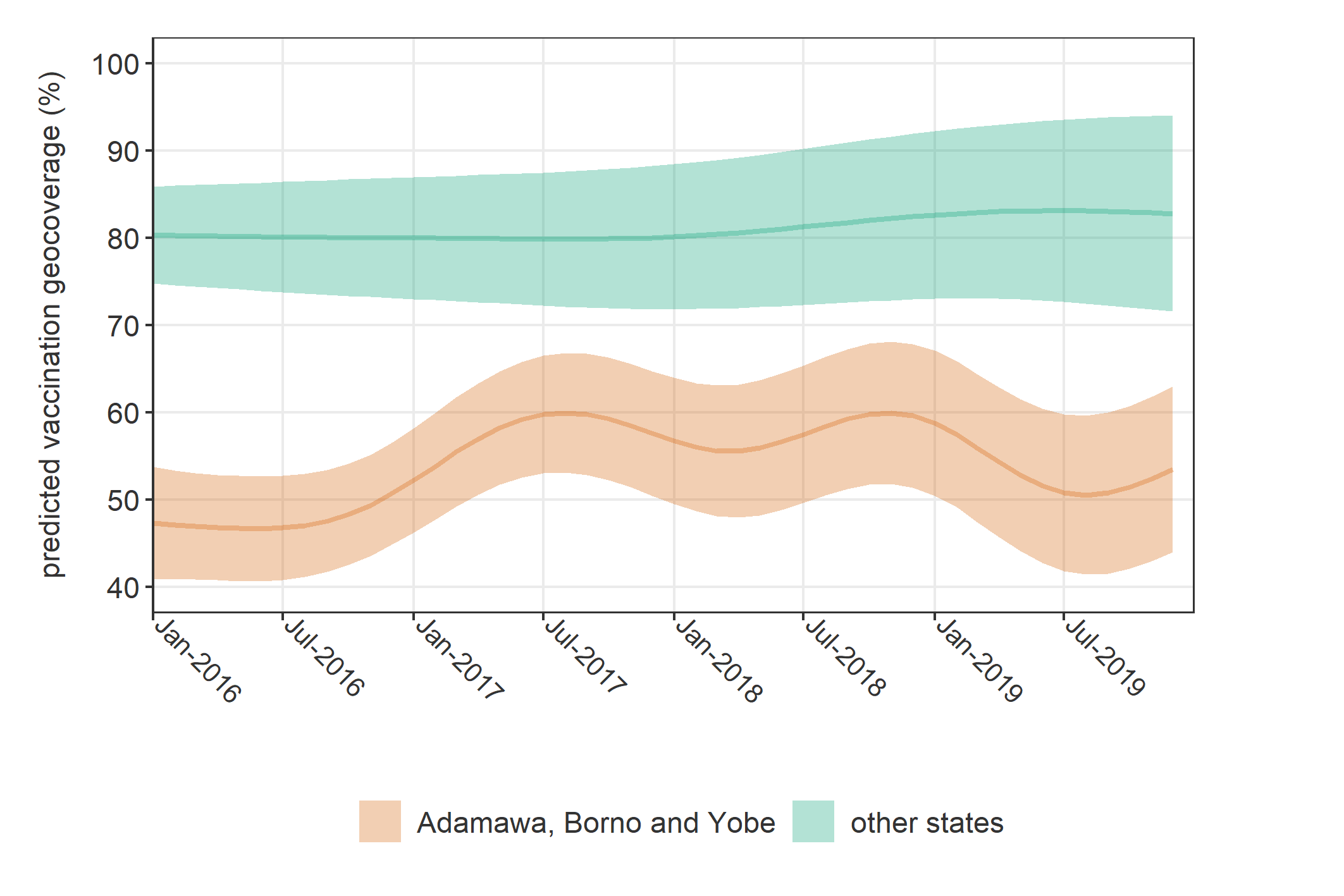

Figure S9. Estimated vaccination geo-coverage as predicted using generalised additive modelling from VTS data, for Adamawa, Borno and Yobe and other states, respectively. Shaded areas represent 95% confidence intervals.

### Results

#### Predictive model

Figure S10 and Figure S11 show the model’s out-of-sample predictive accuracy on cross-validation. In practice, both graphs represent a worst-case scenario in which prediction always takes place in LGAs that the model has not been trained on (i.e. new levels of the random effect). Because survey coverage in Nigeria is very high, in reality both models predict deaths mostly for ‘within-sample’ LGAs.

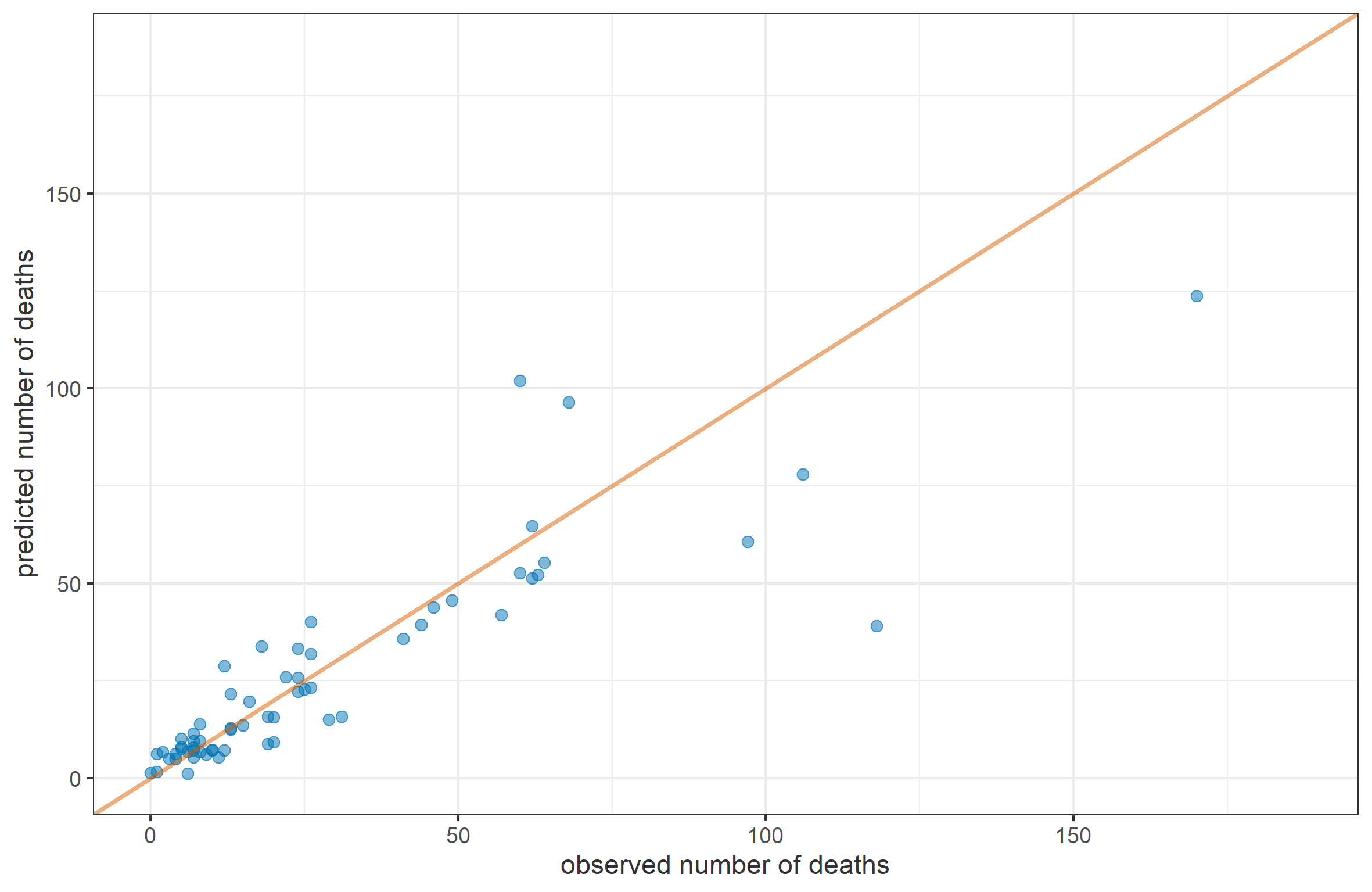

Figure S10. Predictive accuracy of the model for CDR on ten-fold cross-validation. Each blue dot shows the number of observed and predicted deaths by LGA. The red line shows perfect prediction.

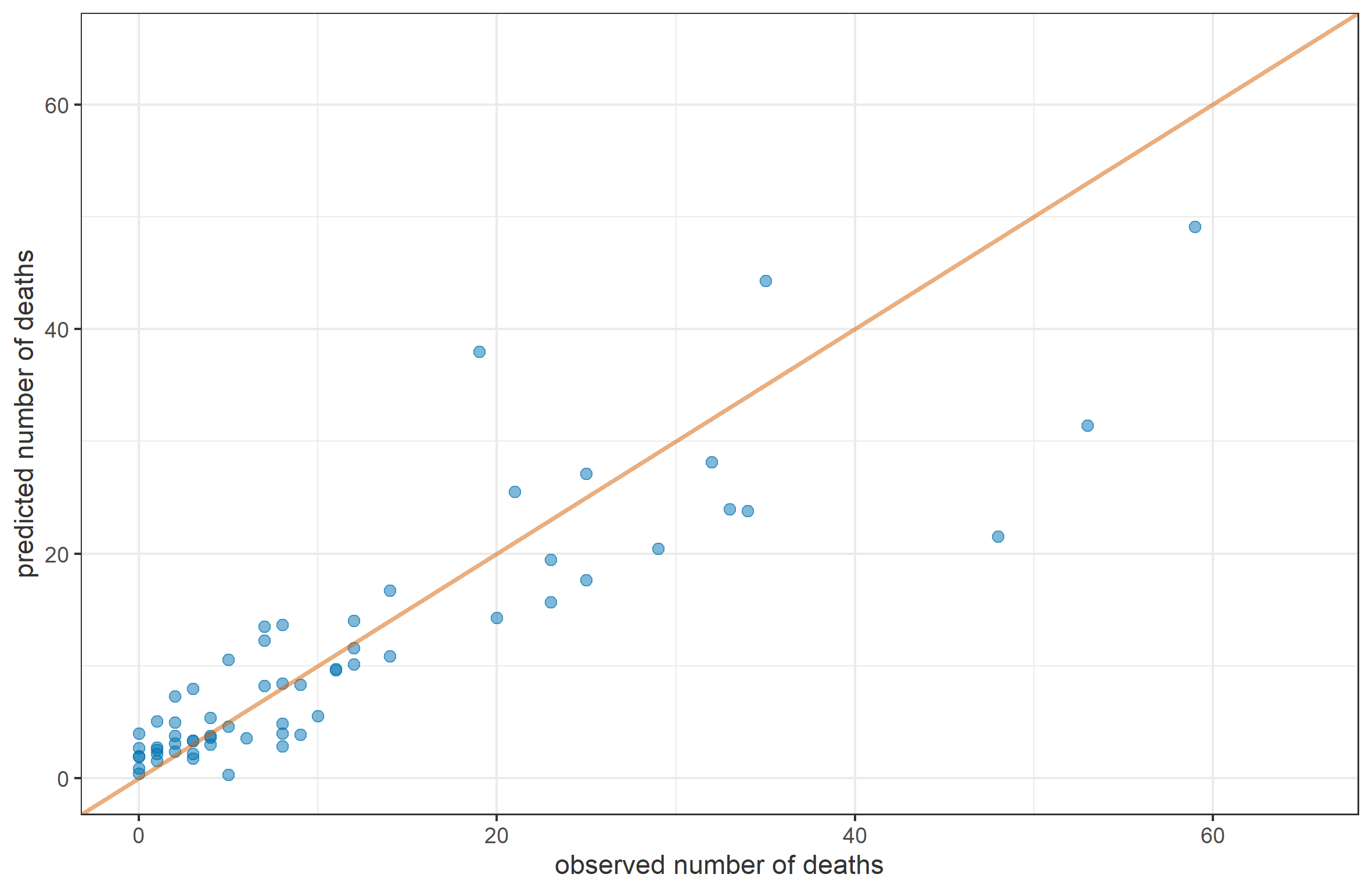

Figure S11. Predictive accuracy of the model for U5DR on ten-fold cross-validation. Each blue dot shows the number of observed and predicted deaths by LGA. The red line shows perfect prediction.

### Discussion

#### Sensitivity analysis: bias in displacement and population data

Figure S12 shows how the central estimate (likely counterfactual scenario) of total and excess death toll varies as a function of potential bias in both the four base population estimates and the number of IDPs as reported in the DTM database.

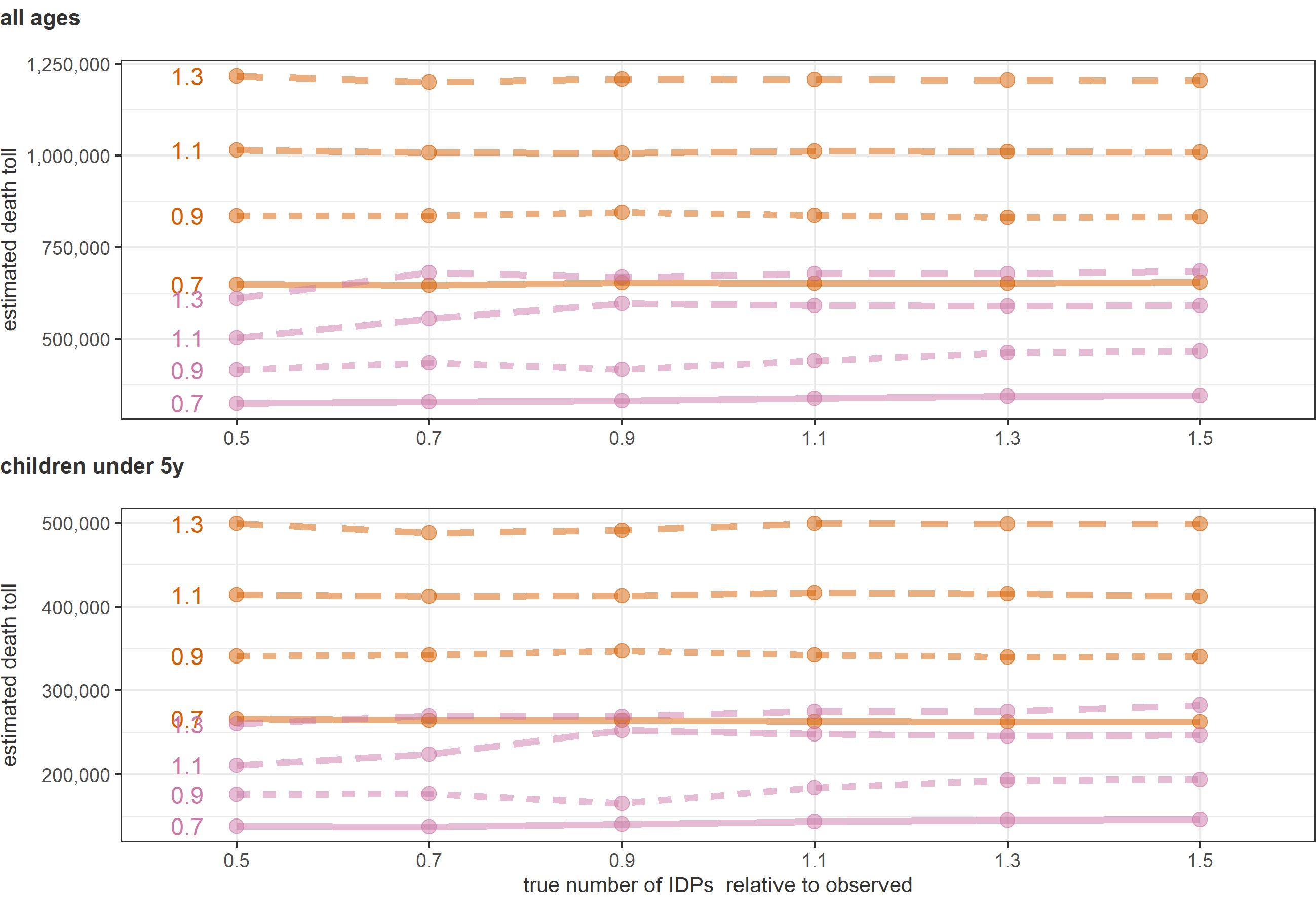

Figure S12. Sensitivity of the estimated total (orange) and excess (pink) death toll (as the point estimate given the most likely counterfactual scenario), for both all ages and children under 5y, to varying levels of bias in input population and IDP data. Each sensitivity value is a ratio of actual to observed values, i.e. multiplier applied to the observed data (accordingly, values < 1 imply under-estimation, and vice versa). Sensitivity values for the ratio of true to observed population figures are indicated within the plots to the left of the corresponding output estimates.

#### Sensitivity analysis: Under-estimation in U5DR

Figure S13 shows how the central estimates of total and excess death tolls, by age group, vary with increasing percentages of under 5y deaths that were not detected during SMART surveys.

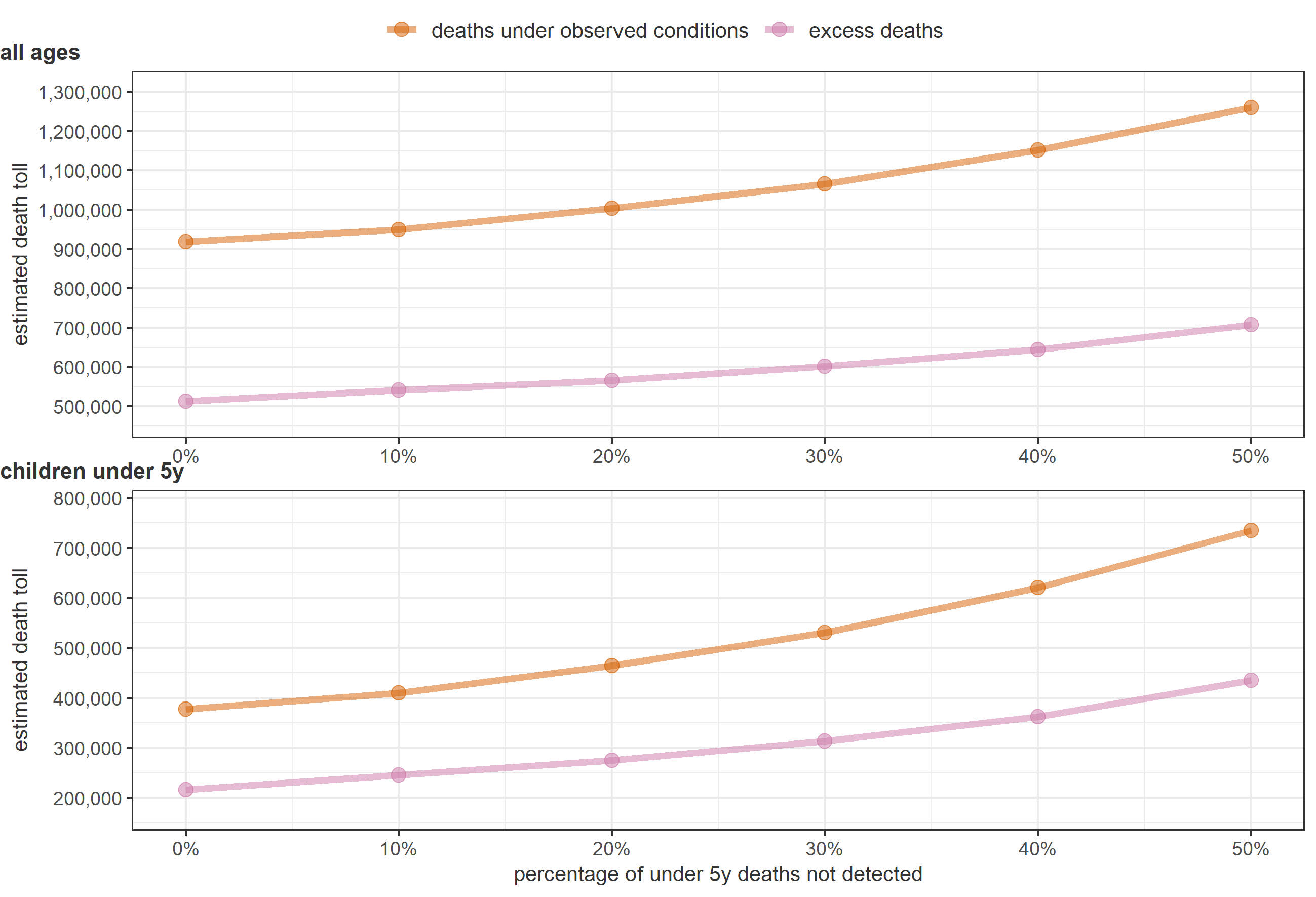

Figure S13. Sensitivity of the estimated total and excess death toll (as point estimates given the most likely counterfactual scenario), for both all ages and children under 5y, to varying levels of bias in input U5DR survey data. Each sensitivity value is a percentage of under 5y deaths that may have been missed during survey interviews.

#### Trends in the rate of people killed, by accessibility status of LGAs

Figure S14 displayed reported rates of people killed (combining the different insecurity monitoring project datasets with our reconstructed population denominators) by source and whether the LGA was fully accessible or not during any given month (accessibility was determined by us based on document review: see Table 1, main text).

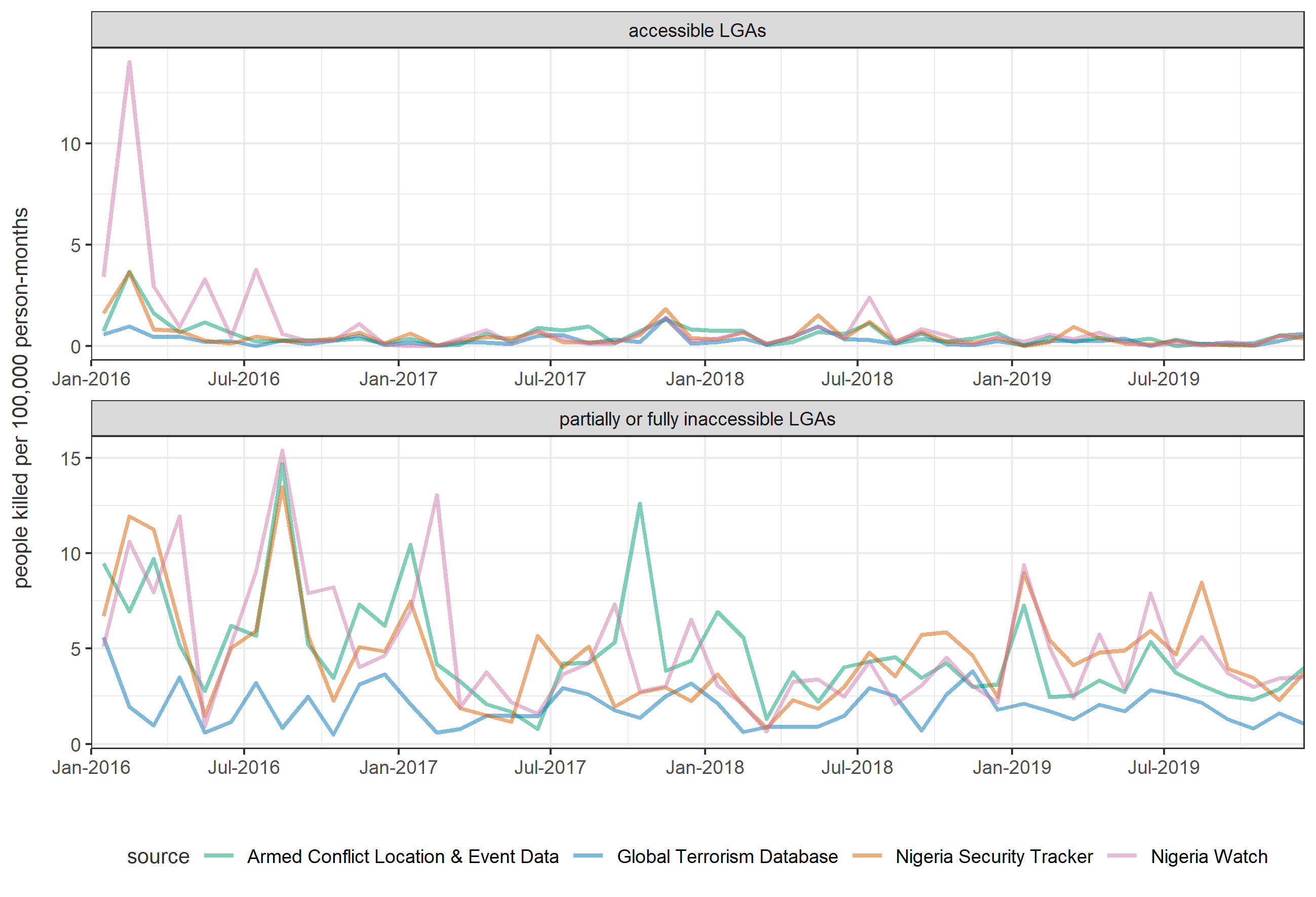

Figure S14. Trends in the monthly rate of people being killed according to each of the four insecurity monitoring projects for which data were available, and by whether LGAs were accessible or partially / fully inaccessible.

#### Sensitivity analysis: potential inaccessibility bias

Figure S15 shows how the total and excess estimated death toll varies if one assumes varying levels of bias in the estimated CDR for LGAs that were partly or completely inaccessible during any given month. As expected, results are highly sensitive to this potential bias in Borno state, where most LGAs were at least partially inaccessible during the analysis period.

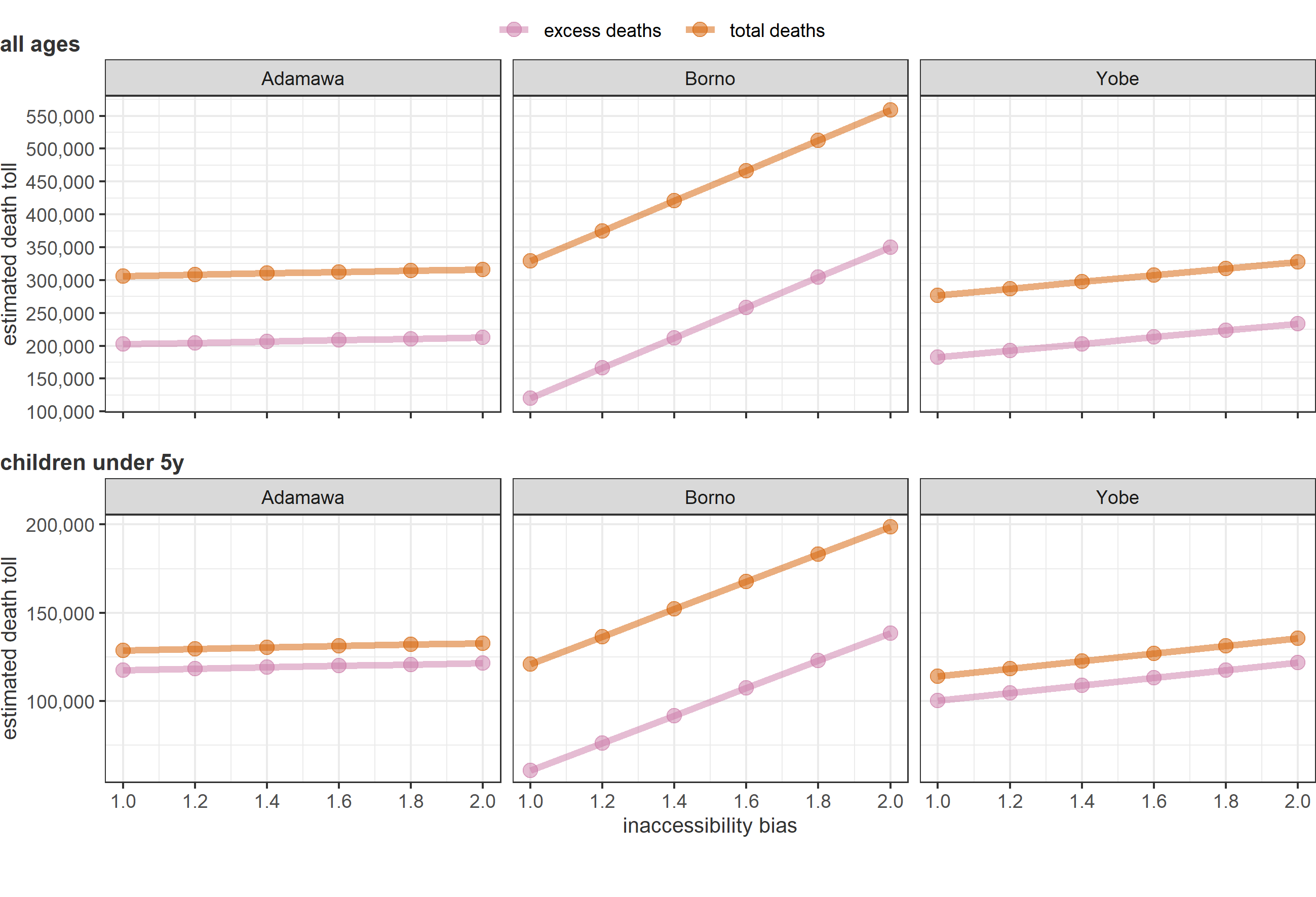

Figure S15. Estimated total and excess death toll, by age group and state, for increasing values of potential inaccessibility bias (each value is a multiplier for the estimated CDR, applied for LGA-months where accessibility was partial or none).

### References

1. Erhardt J. Emergency Nutrition Assessment (ENA) Software for SMART. 2020. https://smartmethodology.org/survey-planning-tools/smart-emergency-nutrition-assessment/.

2. Weber EM, Seaman VY, Stewart RN, Bird TJ, Tatem AJ, McKee JJ, et al. Census-independent population mapping in northern Nigeria. Remote Sensing of Environment. 2018;204:786–98. doi:10.1016/j.rse.2017.09.024.

3. Methodology: High Resolution Population Density Maps + Demographic Estimates. Facebook Data for Good. https://dataforgood.fb.com/docs/methodology-high-resolution-population-density-maps-demographic-estimates/. Accessed 23 May 2021.

4. Higgins J, Adamu U, Adewara K, Aladeshawe A, Aregay A, Barau I, et al. Finding inhabited settlements and tracking vaccination progress: the application of satellite imagery analysis to guide the immunization response to confirmation of previously-undetected, ongoing endemic wild poliovirus transmission in Borno State, Nigeria. Int J Health Geogr. 2019;18:11–11. doi:10.1186/s12942-019-0175-y.

5. Abdelmagid N, Checchi F. Estimation of population denominators for the humanitarian health sector: Guidance for humanitarian coordination mechanisms. London: London School of Hygiene and Tropical Medicine; 2018. https://www.who.int/entity/health-cluster/resources/publications/LSHTM_Population_Guidance_GHC_15-Nov-2018.pdf. Accessed 21 May 2021.

6. Buuren S van, Groothuis-Oudshoorn K. mice: Multivariate Imputation by Chained Equations in R. Journal of Statistical Software. 2011;45:1–67. https://www.jstatsoft.org/v45/i03/.
